# Plaintiff experiences of the medico-legal environment in Ireland

**DOI:** 10.1101/2024.09.16.24310069

**Authors:** Suzanne McCarthy, Mary Donnelly, Aislinn Joy, Elaine Lehane, Peter O’Sullivan, Eimear Spain

**Affiliations:** School of Pharmacy, University College Cork; School of Law, University College Cork; School of Medicine, University College Cork; School of Nursing and Midwifery, University College Cork; School of Law / Faculty of Education and Health Sciences, University of Limerick

## Abstract

**Introduction:** The recent surge in clinical claims in Ireland has sparked concerns about the unsustainable trajectory of medical negligence litigation. Current evaluations have primarily focused on the financial and temporal aspects of litigation, leaving a gap in understanding the experiences of plaintiffs within the adversarial system. This study aims to fill this gap by critically exploring the experiences of those affected by medical negligence and the ensuing legal process.

**Methods:** A qualitative descriptive approach was employed to explore the experiences of plaintiffs following patient safety incidents and their interactions with the legal process. Semi-structured, open-ended interviews were conducted with participants who had been involved in medical negligence litigation in Ireland. Participants were eligible for inclusion if they were aged 18 years or over and were involved in medical negligence litigation in Ireland (as a plaintiff), whether the case was resolved by negotiated settlement, a form of alternative dispute resolution (e.g. mediation), or trial hearing. Maximum variation sampling was used to capture a diverse range of experiences, with sample size determined by the concept of ‘information power.’ Recruitment was facilitated by the Health Service Executive (HSE) through invitations sent by The National Open Disclosure Office. Interviews were conducted in-person or online, recorded, transcribed, and analysed thematically. Ethical approval was obtained from the Social Research Ethics Committee of University College Cork.

**Findings:** This research presents the views and experiences of fifteen participants; eleven participants shared their experiences relating to an adverse event which impacted a family member (one participant spoke about two family members), nine of whom were children (including both minors and adult children), and three were a spouse. Of the twelve individuals discussed, eight were deceased. Five main themes were identified from the analysis: i) Navigating the aftermath of a patient safety event: Communication, Support and Abandonment; ii) The pathway from adverse event to litigation; iii) Experiences of the Legal System; iv) Emotional and Mental Health Impact of Litigation on Plaintiffs; v) Advocating for Change: Participant Recommendations.

**Discussion:** This research highlights the profound impact of actions taken after a patient safety event on patients, families, healthcare professionals, and organisations, and the importance of Open Disclosure in meeting ethical obligations and ensuring healthcare accountability. It explores the complex relationships between financial compensation, justice-seeking, and the healthcare and legal systems. The findings contribute significant insights to the discourse on medical negligence in Ireland.

## 1.1 Introduction

The recent increase in clinical claims in Ireland has been described as an *“unsustainable trajectory*” with a reported €3.85 billion estimated outstanding required to manage and settle clinical claims at the end of 2022. [1] To-date, evaluation of the dynamic of medical negligence disputes in Ireland has primarily focused on the expediency and costs associated with this type of litigation. Consequently, debates about reform in Ireland have centered on these issues. Whilst the financial and temporal efficiency of medical negligence litigation is important, evaluation of these concerns fails to provide insight into what the adversarial system delivers to litigants, and their experiences of the medico-legal environment. While there have been anecdotal self-accounts in the literature of patient lived views, [2] or media outlets describing the adversarial legal process as “abusive towards patients”, [3] there is an absence of comprehensive data ascertaining the views of these who were subject to the initial iatrogenic harm and the subsequently onerous legal process.

The aim of this research is to address this gap through a critical exploration of plaintiff experiences with the medico-legal environment.

To meet this aim, a review of the literature was undertaken and is described in section 1.2. Following this in section 1.3, we present the findings of a qualitative interview study. In section 1.4, we triangulate the findings presented in 1.2 and 1.3 to develop a more comprehensive understanding of the phenomenon.

## 1.2 Literature Review

### 1.2.1 Background

The aim of this literature review is to review empirical literature both in Ireland and internationally, where patients involved in medical negligence were the subject cohort of the study. There is an emphasis on alternatives to the current tort-based system, such as no-fault liability, [4] or alternative dispute resolution [5] in this literature. While neither of these have gained favour in this jurisdiction to date, review of these international studies may provide insights into patients’ needs, opinions and experiences in legal systems which have entertained such reforms.

Furthermore, there is also a growing body of academic literature emphasising the psychological damage that adverse events and associated litigation can have on patients and their families, and the importance of placing patients’ emotional needs to the forefront of any recommendations in this future. Iedema notes that “*What none of the data sources….will illuminate however is what healthcare-caused harm means to patients, and how the complex consequences of such harm for patients and families are to be understood and tackled.”* [6]

### 1.2.2 Methods

Traditional database searching was employed on *PubMed* and *Google Scholar* using a combination of the following terms: *“medical negligence”* OR *“clinical negligence”* OR *“malpractice”* AND *“reform”.* While these searches may provide a myriad of irrelevant results it was found that limiting the search by the inclusion of phrases such as “qualitative” or “interview” or “patient experience” was too restrictive and in fact may result in seminal papers being excluded. General searching on Google, governmental websites and legal databases, e.g., *Westlaw* (IE & UK) and *Lexis* was also carried out. The citations of certain key papers were also examined.

The paucity of research detailing participants’ views became very apparent early on. Some of this may be credited to several barriers, such as a reluctance on the part of organisations to release information regarding matters of a “protracted and confidential nature”. [7] Equally conceivable is the contention that patients who have endured medical injuries, with the subsequent adversarial legal proceedings, may be unwilling to revisit such events for research purpose, so as not to re-live the emotional anguish they initial suffered from their ordeal. [8][9] Another concern noted in one study was that such research may entice patients of settled cases to initiate legal proceedings. [9]

While many of the studies uncovered interviewed an array of parties, it is important to remember that the primary focus of this research was on the views of the patient or plaintiff. Other studies have explored the views of physicians and risk managers, and legal professionals.

### 1.2.3 Findings

| Study Title | Publication Year | Category of Participants | Jurisdiction | Study Design |
| --- | --- | --- | --- | --- |
| *Plaintiff aims in medical negligence disputes: limitations of an adversarial system* [10]. | 2023 | • Barristers (n=12)<br>• Patient representative groups (n=2) | Ireland | Semi-structured interviews |
| *Medical Dispute Committees in the Netherlands: a qualitative study of patient expectations and experiences* [11]. | 2022 | • Patients/family members (n=26) | Netherlands | Qualitative semi-structured |
| *The value of compensation* [12]. | 2022 | • Patients (n=15) | United Kingdom | Qualitative interviews |
| *The struggle against perceived negligence. A qualitative study of patients' experiences of adverse events in Norwegian hospitals [13].* | 2018 | • Patients (n=15) | Norway | Qualitative interviews with explorative descriptive design. |
| *Improving reconciliation following medical injury: a qualitative study of responses to patient safety incidents in New Zealand [9].* | 2017 | • Patients (n=62)<br>• Hospital Administrators (n=12)<br>• Lawyers (n=5)<br>• Accident Compensation Corporation (ACC) staff (n=3) | New Zealand | Semi-structured interviews |
| *A Study of Medical Negligence Claiming in Scotland: Research Findings [14].* | 2012 | • Patients (n=32);<br>• 42 - other stakeholders (i.e. solicitors, support groups and medical practitioners) | Scotland | Interviews with patients and focus groups with stakeholders. |
| *The Impact of Disclosure of Adverse Events on Litigation and Settlement. A review for Canadian Patient Safety Institute. (CPSI Study) [15].* | 2007 | • Patients/families (n=11);<br>• Risk management/insurance providers (n=8);<br>• Legal counsel (n=6) | Canada | Informal Survey consisting of open-ended questions |
| *Why do people sue doctors? A study of patients and relatives taking legal action (Lancet Study) [16].* | 1994 | • Patients (n=227) | United Kingdom | Survey |

### 1.2.4 Discussion

#### The *Status Quo* and Reform Proposals for Irish Medical Negligence Law

In Ireland the adversarial court system which facilitates plaintiffs to claim damages for medical negligence is based on the tort principle of *duty of care* and the standards which a doctor must have fallen short of to be liable for negligence are enunciated in *Dunne v National Maternity Hospital*:

> “[w]hether he has been proved to be guilty of such failure as no medical practitioner of equal specialist or general status and skill would be guilty of if acting with ordinary care…etc…” (per Finlay CJ). [17]

There are of course instances where the judiciary, mindful of potentially opening the floodgates for compensation, has implemented demarcation lines in this jurisprudence. In *Byrne v Ryan*, it was held that a mother seeking compensation for the upbringing of two healthy children following a failed sterilisation would not be a *“fair and reasonable”* request to impose on the defendant.[18]

There have been reform proposals put forward by various review groups in Ireland, but a detailed discussion is beyond the scope of this study, as many did not encompass patient groups in their data. The recent Department of Health Expert Report Group considered the introduction of no-fault compensation,[19] which is currently present in France, New Zealand & Scandinavian countries (see *infra* Diagram 1). It deemed such a scheme to be unconstitutional in Ireland, as it would deny patients the right to access to the courts [19]. The Group also did not favour the introduction of a Medical Injuries Assessment Board (MIAB) as a mechanism to streamline disputes akin to that already seen in personal injury disputes. This distinction was due to the inherently complex nature of medical disputes, and the requirement for specialised expert evidence in each case, which would not lend itself to a paper-based book of quantum for assessing claims.[19]

**Diagram 1:**
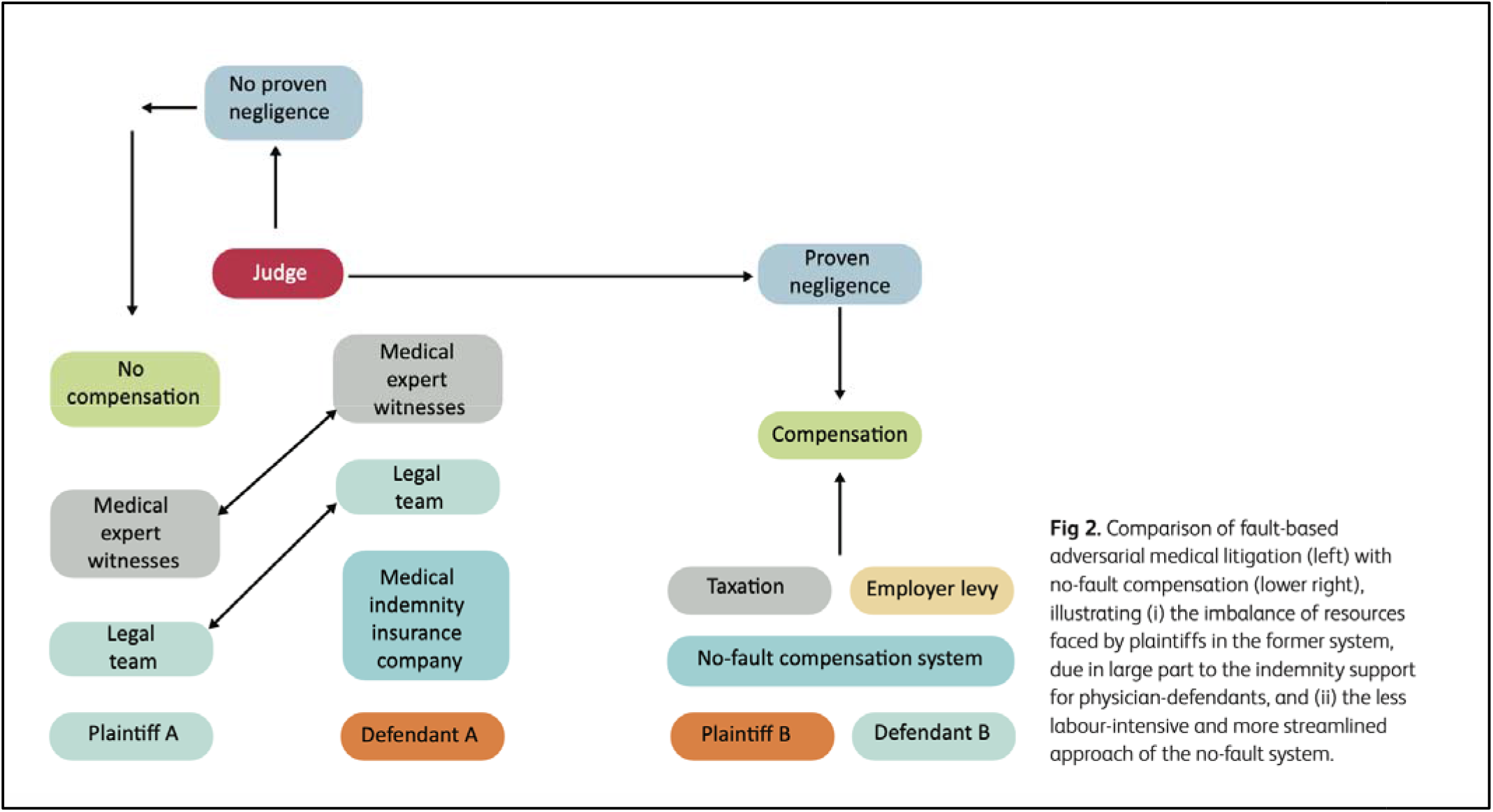
Comparison of fault-based adversarial system v no-fault compensation (*per* Epstein R 2023) [20].

#### Emerging Narratives from international studies regarding patient perspectives

##### Irish 2023 Study: [10]

The author of this piece notes that research in Ireland regarding plaintiff aims, expectations and experiences is *“non-existent”*, with the primary focus of the academic debate centred on tort-based compensation reform (see *supra* discussion of Expert Group)[19], without due consideration to the patient themselves. This study interviewed barristers and patient representative groups, and divided what they considered to be patient aims into the following two subheadings:

1. Aims that are achievable within the Irish adversarial system (“*realistic*”), i.e. financial compensation and a day in court, and
2. Aims that are catered for by settlement proceedings (*“extralegal”*), i.e. an explanation, an apology and prevention of recurrence.

In terms of financial compensation, the severity of the illness was seen to play a significant role, and for those with severe injuries, an immediate need to ensure the patient is looked after for the rest of his/her life was evident. It is also noted by many that to endure the “uphill” battle of protracted legal proceedings, compensation is not the sole motivation, and many are anxious to prove categorically that they were wronged.[10] It was found that a minority of clients would want a day in court, perhaps as an opportunity to be heard in an unequivocal manner. The majority are content with settling their case, and this is borne out in statistics from the NTMA, with 98% of cases being settled. [21] The author suggests another reason for plaintiffs opting for this approach may be due to legal conditioning by their solicitor who would persuade them to not pursue their case as a more realistic and attainable option would be to settle. [22] An alternative venue such as mediation may be a more appropriate forum for plaintiffs to have their views heard in a non-adversarial manner, however, the uptake of this approach to date in Ireland has been quite low. [10][5]

In terms of obtaining explanations, the author notes that there is a disconnect between policy and practice; although open disclosure may be embedded in HSE policy and legal protections provided for those who engage in open disclosure in the Civil Liability Amendment Act 2017, it is often not forthcoming for fear of litigation. For instance, one representative from a patient support group membership states:

> “We talk about open disclosure as a policy, policies are all very fine but if the people on the ground aren’t operating out of those policies, well then they count for naught.” [10]

The remaining extralegal aims are primarily related to apology, which may be potentially beneficial to the patient in terms of their emotional healing. This is something that the current Irish tort system cannot cater for, nor the desire to prevent recurrence.

This author has also written extensively on the topic of this and has detailed the emotional and psychological burdens medical negligence litigation can have on patients [8] and practitioners (“the second victim”) alike.[23]. Any reform proposals in Ireland should be cognisant of these shortcomings, which have not been critically reviewed at a governmental level to date. It is also noteworthy that these studies did not include patient populations in the study cohort, beyond the patient representative groups, but as they were extremely pertinent to the Irish approach, they were included in this literature review.

##### Dutch 2022 Study: [11]

Since 2016 a new approach to medical negligence in the form of *dispute committees* has been adopted in Holland as a “hybrid” independent forum to provide monetary compensation, and an affirmative verdict without protracted legal proceedings in a courtroom. [24] A key expectation for taking cases regardless of the verdict was a desire to be *heard* and make *positive improvements* to the healthcare system. Some participants reported feeling *silenced* due to the time constraints of the proceedings, i.e. requirement to summarise your complaint in 3 main points in under 10 minutes. For most participants financial compensation was a priority (claims capped at €25,000), for others it was “the last thing on their mind” and feelings of retribution weighed more heavily. There were suggestions of an *“unequal power relationship”* for participants taking a case, due to a variety of considerations including the formality of the committee or the complicated nature of filing a complaint. The absence of the doctor, who was responsible for the hearing, was a source of contention for many participants as they felt a substitute representative was not equivalent. [11]

One key issue that was raised was the lack of impartiality in the complaint committee, with some members sitting on internal complaint committees in the hospital, as well as on the independent led committee. This resulted in an impression of pre-determined decisions: *“I felt as if the verdict was already sitting in the top drawer. The president was not interested in anything.”*[11]

The authors summarised some of the grievances felt by participants as also being a product of epistemic injustice, which arise essentially due to the invidious position of simply being a patient/plaintiff, namely (1) they are not as well-resourced in medical/legal knowledge or as financially equipped as the defendant (*hermeneutical injustice -* prejudices due to structures), and (2) the many stereotypes of being weaker both physically and mentally, perhaps result in less credible testimonies in proceedings (*testimonial injustice*). Throughout the dispute committees many instances of this were seen, e.g. patients not being heard, and difficulty processing the legal and medical jargon employed in such situations, without the need to enlist the support of a legal representative.[11]

##### United Kingdom 2022: [12]

This study, conducted by Opinium, demonstrated that compensation is *“not life changing”*, but a step in the process of patients rehabilitating their lives and reassuring them for the future for both medical bills and any potential loss of earnings. The study spoke of the two facets of medical negligence cases, firstly the physical disability, and secondly the mental health component. Participants spoke of feelings of isolation, anxiety, low mood, a lack of confidence, and post-traumatic stress disorder. [12] One mother who lost a son from a preventable disease (undiagnosed Addison’s disease) spoke of it fracturing her relationship with her remaining children, and her PTSD causing her to be suicidal. The manner in which an apology was given varied between interviewees. Some were provided with a clear, transparent acknowledgement of wrongdoing, and recommended the patient seek legal action to compensate for the future. However, the reality for many others was a lack of any form of empathy or compassion, and in fact attempts to divert blame from the NHS to the patient were put forward, which shifted a huge amount of the legal responsibility of claiming onto the patient. This was seen as prolonging an already arduous and complex legal proceeding: *“They basically said that I was lying or embellishing [it]”.* [12]

##### Norwegian 2018 Study: [13]

Norway and other Scandinavian countries had in operation a taxation-funded compensation scheme akin to the no-fault based model as outlined above since the 1980s.[25] In this study the experiences of hospital patients following orthopaedic, surgical or oncology-based adverse events were examined. It is interesting to note that during the recruitment phase, 60 invitation letters detailing the study were send out, from which 19 responses were obtained, but some opted not to participate as they “*had enough trouble*” caused by the event in question and others were excluded from the study as they were unable to focus clearly during the interview. [13] This is something worthwhile for researchers to be mindful of with this particular cohort of interviewees. Another important parameter is that not all participants had claimed compensation from the district’s respective injuries ombudsman.[13]

Many of the patients accepted that in all hospital practices there is an element of *risk*, however they felt that when negligence did become clearly evident, the hospital staff engaged in defensive and *“cover-up”* practices and were not forthcoming towards the patient in providing explanations, support or in some instances refusing follow-up care. This left many patients with an impression that their wellbeing and health was being severely undervalued by the hospital, e.g. reasons for delays and mistakes in diagnosis not being properly explained and sometimes in fact being trivialised as *part and parcel* of the recover process:

> “At home, I suddenly could not hold my arm out. I contacted the doctor at the hospital; he said it was completely normal and I could relax. It would get better again. But it certainly did not: it got worse and worse. I spoke to the hospital again but got the same answer …”[13]

Patients who were facilitated with disclosure proceedings often did not obtain an apology or a detailed explanation, and oftentimes the practitioner in question was either not present or genuine in their response. This again was seen by many as marginalising their feelings, and not conducive to their aims. While the article did not detail compensation *per se,* it did allude to allied staff members (e.g. nurses and physiotherapists) recommending patients to avail of this service. The authors noted limitations in the sample size (n=15) for the general applicability of their findings, and they suggested that a potential rationale for the hospital’s normalisation of negligent practice may be due to a culture of collegiality or efforts to avoid reputational damage for the hospital. This study is a stark reminder of the importance of patient inclusion from the initial error through to any subsequent disclosure meeting. [13] This is a sentiment echoed in much of the academic literature, which emphasises openness as a requirement for patient healing, safety and learning. [26].

##### New Zealand 2017 Study: [9]

In New Zealand a non-litigious approach (i.e. no-fault compensation) is taken to medical negligence cases,[20] where the state-funded Accident Compensation Corporation (ACC) provides patients with compensation and reconciliation options, which removes the necessity for plaintiffs to pursue legal options. [27] While this approach is the complete antithesis to the Irish adversarial setting, it is nonetheless an informative study and still shows that patients’ experience even in reformed legal systems have unfulfilled aims. [9]

This study advocated for a tailored patient-centric approach to the reconciliation process by the organisation and physician deemed responsible for the patient safety incident. The following were identified as salient themes for the patients:

- Being *asked* in a clear and comprehensive manner what remedial action is required for them. This is important because all patients require individualised responses to the unique distress and not a simple homogeneous one, e.g. a memorial plaque hung in the hospital.
- Being *heard* is something the authors noted as being novel to the literature, with only 27% of participants feeling that they were appropriately listened to and not interrupted during the reconciliation process. Many considered this process to be therapeutic for the patient’s emotional needs to recount their entire story with the physician in audience:

> “It was that [being heard and acknowledged during reconciliation meetings] not the compensation, that made me feel healed. It restored my trust in my doctor.”

- A genuine *apology* offered to the patient and executed in a time appropriate manner, i.e. when best suits the patient and not months after the incident occurred. The inclusion of a request for forgiveness from the clinician was also welcomed by participants.
- Appropriate *terminology* used - patients took particular umbrage to the use of the word *“resolution”,* as it gave the impression that this matter was completed, but in actual fact the emotional and life altering consequences of the injury suffered are never truly “resolved”. Alternative nomenclature proffered included “reconciliation” or an “ability to move on if the provider responds appropriately”.
- *Support personnel* in the room with the patients, e.g. lawyers or family members. [9]

This study provides insights that physicians and hospitals should be adaptive and reactive to the patient’s emotional needs in the aftermath of a medical negligence incidents, and this was consistent with theoretical literature relating to therapeutic jurisprudence (i.e. the philosophical study underpinning how the legal process can produce psychological benefits or detriments to litigants). [28]. This was particularly evident with the value placed on *being heard* during the reconciliation process.

##### Scottish 2012 Study: [14]

This study relates to compensation reform proposals in Scotland for medical negligence cases, namely the introduction of a no-fault programme and its viability in catering for patient needs.[20] The main findings pertaining to patient views related to a lack of proper communication or explanation during the complaint process, and a desire to safeguard future patients from similar incidents. Again, as in similar studies, patients stated that financial reward was not the main rationale for pursuing litigation. While the authors saw potential patient benefits from a capped compensation scheme in overcoming certain legal barriers (no longer required to prove the standard of negligence or pay legal representation fees in litigation), it was conceded that institutional reform would also be necessary.[14] Cave notes that the findings of this study while being informative, would not be directly transferable as a reform proposal for the United Kingdom as a whole, given the vast population, and “the strains it would put on the NHS”, particularly in the short to medium term. [29]

##### Canadian 2007 CPSI Study:[15]

In this survey carried out by the Canadian Patient Safety Institute (CPSI), patient participants did not consider monetary compensation to be the primary motivator for instigating litigation, but it was beneficial to garnering the attention of the organisation or healthcare provider and forcing them to take responsibility for the alleged claims. They stated there was three reasons that motivated litigation - (1) obtain information; (2) an acceptance from the organisation that an error was the causative factor (*accountability*); and have a resolution to the manner they were treated following the adverse event:

> “We had worked for years having meetings with [medical directors and senior executives] before they even acknowledged a problem - by then it was too late.”

In terms of disclosure, patients considered it an essential part of the process, and it must be *“compassionate, patient-centred”* and encompass an apology. Some even considered it to potentially reduce the frequency of litigation, as it would cater for patients’ needs for emotional redress, explanations and an apology. This is in contrast to the literature findings, where it was accepted that a complete absence of disclosure or apology would have a probable chance of increased litigation. [30] In this instance, the determining factor for patients’ decision to pursue financial compensation would be predicated on the category of harm caused (i.e. *mild*, *moderate* or *severe*), regardless of open disclosure protocols being adhered to or not. This view would correlate with that of the legal and insurance representatives interviewed. [15]

A gap in the empirical literature was noted by the authors of this study in that there is no material involving the motivations of patients who opted not to seek compensation or initiate a complaint in a disciplinary forum. This view is echoed by Mazor *et al.* noting that:

“*It would be very informative to query patients who believe they have been harmed but decided not to sue”.* [31]

##### United Kingdom 1994 Lancet Study:[16]

In this Lancet study, 70% of the patients who agreed to take part in this study were living with long-term effects as a result of severe harm caused, which impacted on their work, social life and family. There were four main motivations identified in this study for the participants willingness to *“endure a long and often frustrating legal process:”*

1. assurances that an appropriate *standard of care* will be complied with for future patients, and lessons learned from their incident;
2. an *explanation* as to how, and why the injury happened to them;
3. the staff and organisation involved held *accountable,* and disciplined if necessary; and
4. financial *compensation* for actual loss and suffering incurred from medical negligence, and any future care necessary.

Secondary issues revealed in the study related to shortcomings in communication subsequent to the adverse event, and even a *“reluctance to apologise or being treated as a neurotic.”* With this in mind, the authors concluded the piece echoing a narrative emphasised in aforementioned studies above, that financial compensation is not the sole resolution that patients seek, and in fact *open* and *transparent* communication is pivotal for them to process the trauma they have endured. This is one of the key reasons the authors considered a *“no-fault compensation system”* to not be satisfactory in resolving all the patient aims, as it looks at the problem in isolation, i.e. exclusively from a financial perspective.[16]

## 1.3 Qualitative Interview Study

### 1.3.1 Introduction

In this section, empirical findings in respect of the plaintiff experience of the medico-legal environment in Ireland are reported. The section begins with an explanation of the methodological choices made and then presents the key findings. In addition, other direct evidence of plaintiff experience in the public domain, found in submissions made to the Expert Group to Review the Law of Torts and the Management of Clinical Negligence Claims as related to the interview study findings are presented. The report concludes with key observations to further inform the discourse of medical negligence in Ireland.

### 1.3.2 Methodological Approach and Methods

We adopted a qualitative description approach to study design. Such an approach is appropriate when engaging with people who have direct experience of a particular phenomenon and is helpful when describing phenomena and highlighting current practices, trends, or patterns. [32] It allows for both ‘description’ of a multifaceted phenomenon in its real-world context but also recognizes and incorporates uncertainty about the phenomena being studied and the research methods used to study them. [33] By rigorous design, conduct and analysis, we sought to answer the following research questions:

i. What are the experiences of plaintiffs following a patient safety incident?
ii. What are the plaintiff experiences of the legal process?

#### Methods

Data were collected through the conduct of semi-structured, open-ended interviews. An interview guide was developed based on the rationale for the study, the key questions of interest and a review of the relevant literature.

Inclusion criteria for participation were aged 18 years or over and involvement in medical negligence litigation in Ireland (as a plaintiff), whether the case was resolved by negotiated settlement, a form of alternative dispute resolution (e.g. mediation), or trial hearing.

Maximum variation sampling was deemed most appropriate so as to include as wide a range of participants and experiences. [34] Sample size determination was predicated on the concept of ‘information power’ which directly relates to the amount of information a sample holds relevant to the study focus and specificity of participant experiences. [35] Recruitment was undertaken with the support of the HSE; a letter of invitation to participate in the research was sent by The National Open Disclosure Office to potentially interested parties including advocacy groups in this area.

An information sheet and consent form were sent to all participants at least 24 hours in advance of the interview. Informed consent was provided by the participants. The participants were informed that they had the right to withdraw from the research at any time within two weeks of the interview. They were also assured that confidentiality would be maintained during the write-up of the research with pseudonyms to be used throughout.

The interviews were conducted by the researchers (ES and MD), and based on the preference of the participant, were conducted in-person or online using Microsoft Teams software. The recording was transferred to UCC OneDrive. The recordings were transcribed, verified, and reviewed by the research team. Thematic analysis was subsequently undertaken [36,37]. A comprehensive thematic summary, which moved beyond individual participant reports by developing an interpretation of a common theme, is presented.

This research received ethical approval from the Social Research Ethics Committee of University College Cork (Ref: 2023-105). A subsequent amendment to extend the time of the project to December 2024 and to allow participants to review the transcript was approved by the Social Research Ethics Committee in October 2023 (Ref: 2023-105 Amendment).

### 1.3.3 Findings

This research presents the views and experiences of fifteen participants consisting of twelve females and three males. Eleven participants shared their experiences relating to an adverse event which impacted a family member (one participant spoke about two family members), nine of whom were children (including both minors and adult children), and three were a spouse. Of the twelve individuals discussed, eight were deceased.

Four participants spoke about their personal experience of an adverse event.

The experiences discussed by participants related to a diverse range of adverse events including misdiagnoses and missed diagnoses, cases related to Cervical Check, neonatal injuries, and surgical complications. The participants also had varying experiences following the adverse event; some pursued legal proceedings and settled prior to trial, one participant described proceeding to trial and some initiated but did not pursue a case. The adverse events and the experiences discussed span from approximately twenty years ago until 2023.

The average duration of the interviews was 48 minutes, ranging from 22 to 70 minutes.

Five main themes were identified from the analysis process:

1. Navigating the aftermath of a patient safety event: Communication, Support and Abandonment
2. The pathway from adverse event to litigation
3. Experiences of the Legal System
4. Emotional and Mental Health Impact of Litigation on Plaintiffs
5. Advocating for Change: Participant Recommendations

#### Theme 1: Navigating the aftermath of a patient safety event: Communication, Support and Abandonment

##### Summary of theme

This theme encapsulates the multifaceted consequences of actions taken in response to patient safety events, with a focus on the intricate dynamics among patients, their families, healthcare professionals, and organisations involved. Within this theme, sub-themes are identified, including deficiencies in communication, discrepancies in disclosure, the transformative nature of interactions with healthcare professionals, the lack of emotional support for patients and families, and a pervasive sense of abandonment. Additionally, the theme recognises the impact of patient safety events on healthcare professionals and acknowledges instances of positive interactions that stand out amidst the challenges.

##### Communication

> “In time of crisis people want to know that you care, more than they care what you know.”

Will Rogers

Effective, compassionate, communication following patient safety incidents is paramount, yet many participants highlighted deficiencies in the communication they received. Some participants faced challenges from the outset, with issues arising during the disclosure of the incident. Participant 4 recounted the lack of compassion and unsatisfactory disclosure of their patient safety incident.

> “everything was just blurted out in black and white, and there you are, and there you go, and off away home with you now, and that’s it. Simple as that. Yes. Go away and live your life now. This kind of stuff. So…it was terrible” [Participant 4]

Some participants described initial positive interactions with healthcare professionals marked by *“initial, honest and humane reactions”*, after which they noted a shift in dynamics during subsequent meetings, where a discernible *“process of damage limitation kicked in” and “it became about protecting the brand”. [Participant 1]*

This was also the experience of Participant 5 who conveyed the transformation in the nature of interactions as healthcare professionals sought to mitigate potential consequences.

> “I thought he [surgeon] was quite a warm person when I met him that night [patient died] like. When we sat crying, I just thought, I felt awfully sorry for him. I felt as sorry for him as I did for myself in that moment. Like I thought, you know, this is just awful to be in this predicament. We’re all in this predicament together and that’s what I felt initially right but that probably changed as time went on. So, at the meeting right, he was very business-like, he was very matter of fact. [Participant 5]

Furthermore, participants experienced a lack of transparency and described the withholding of crucial information in the aftermath of the patient safety incident. Participant 2 described discrepancies between explanations provided by the hospital and observed realities, leading to a sense of manipulation and mistrust. Similar challenges were faced by Participant 7, who recounted similar practices surrounding the adverse event involving their child.

> “there was an explanation, but we knew it was all lies….. It didn’t correspond with what we had clearly seen… There was, there was kind of support, Uh, but it was all an attempt at manipulation. We had some level of engagement umm, but we realized that it was all they were, all lies… to put us off the scent. and the thing was that they weren’t even very good lies” [Participant 2]

> “So there was information held and lies told to us in Hope’s case ” [Participant 7]

Participants also referred to the manner in which information was conveyed to them, both in terms of the verbal and the non-verbal cues, and the impact this had on them. Participant 11 highlighted how the use of medical jargon led to confusion and lack of understanding of the severity of their child’s condition.

> “you could see the, what was the word they used, the white, which is basically if they said words that I understood like and I’m an educated person and if they said words like brain damage instead they used sort of weird words that I don’t understand” [Participant 11]

Participant 12 perceived the way in which she was communicated with and about insulting and one of the motivating reasons for her to pursue litigation

> “ So there were things I felt were definitely you just swept under the rug. But the actual most damning thing I found about them, like I said to you, was the language *they used when they spoke about me or wrote about me. And that’s a thing across the health services. But that’s why I decided I was going to pursue it.” [Participant 12]*

Participant 2 referred to communication they received from the hospital in response to the death of their child.

> “In fact, they did give us a letter basically brushing us off and … the letter was handed to us by the chief executive of the hospital. At the bottom of the letter was a circular mark from a mug of tea. That was on the letter and we looked at this and it was, it was unbelievable. It indicated a level of contempt that was just unacceptable.” [Participant 2]

Participant 1 recalled an encounter with a medical professional who had looked after her son,

> “And I said “while you were talking to me outside Kevin’s room, you were you had your shoulder up against the frame of the door and you had your glasses in your hand and you were twirling them while you were talking to me.”

> I said “that’s appalling, appalling body language when you’re dealing with somebody who’s very concerned.” [Participant 1]

The complex interplay of personal circumstances and perceived power dynamics in the participants’ experiences with healthcare professionals in the aftermath of patient safety events was highlighted by several participants. The feeling of being taken advantage of during a vulnerable time was expressed by one participant who remarked,

> "we just felt that, to take advantage of grieving parents to try and manipulate and lie, at a time like that rather than showing some level of awareness or understanding or sympathy." [Participant 2]

Participant 1 expressed frustration at what she saw as attempts to unsettle her rather than providing answers and support.

> “his [Consultant] first comment was “I believe you have questions - Well, I have questions too”. There was no indication of I’d like to offer you my condolences, which isn’t an admission of anything. There was no humanity, but there was the pushback to put you on a wrong footing and unsettle you. By then I had enough of this man and his atrocious behaviour and attempts to take power.” [Participant 1]

Amidst the participants’ negative experiences with different healthcare professionals, hospital management, and the HSE, there were instances of positive interactions that highlighted the potential beneficial impact of empathetic communication.

> “Dr. X he was extremely helpful and extremely good to us. To the extent that we had his even… he had given us his private phone number and everything. And… you know, we gave him a tough time as well, especially initially before we could… before we knew we could trust him” [Participant 7]

> “She [nurse] was sitting at the side of the bed with me and she was rubbing my arm and she was crying every bit as freely as I was and I was really surprised at that, because after all, we were in ICU” [Participant 1]

> “It depends on the person that you’re sitting across from, because one day you could have somebody who is just not having a good day and no matter what you say to that person they literally don’t care, and then you can meet somebody who is so in love with their job that they will bend over backwards for anybody, and when you talk to them it feels like they’re listening to you. And it feels…it’s actually very emotional when you get somebody like that, because you feel like you want to cry, because somebody is literally looking at you and they have both ears open, and they are taking every single word in, and they feel for you. And it’s a thing of… “Ok, ok, I hear every word you’re saying, leave it with me and I’ll see what I can actually do for you”” [Participant 10]

##### Support

Participants in the study conveyed the absence of emotional, practical and financial support, from healthcare professionals and/or the healthcare system and revealed a notable failure to address the needs of individuals and their families following a serious patient safety event. The absence of emotional support following an adverse event was a consistent theme in this research. One participant whose son had a birth injury described her experience at her 6- week check-up and recalled an absence of emotional acknowledgement or offer of support

> “I remember six weeks I got, I went for a 6 week check afterwards. No sense of listen, you’ve had a big trauma, do you need a bit of support? A bit of help? Do you know how bad it was?” [Participant 11]

Another participant discussing the aftermath of a meeting regarding the Cervical Check audit expressed profound disappointment at the absence of follow-up or psychological support

> “And I was have to digest that news with absolutely nothing. That was that. No, no, no follow up calls to see if you were OK. No psychological support. Absolutely sweet f*** all…It was the most inhumane OK thing I’ve ever experienced; I didn’t even know could exist” [Participant 3]

Furthermore, participants revealed the frustration of having to actively pursue promised supports

> “you’re given all this kind of information, oh people will be in touch with you, we’ll give you this, we’ll give you that, we’ll… a liaison officer will be in touch with you. But then I had to ring four or five times. I said, “Somebody is supposed to be ringing me” and, you know, so I had to follow it up. You know? I had to follow it up… You know, there was, they talk the talk but they don’t walk the walk, you know? On paper it looks like it’s…on paper it looks like that they were doing everything right. “Oh we did this and we had this in place, and we had this in place”. But all of those things, and none of them were connecting.” [Participant 4]

These accounts collectively illustrate a systematic failure in providing the necessary emotional support to patients and their families facing the repercussions of serious patient safety events.

Participants also discussed the absence of financial and practical support in the aftermath of the patient safety incident and their need for more medical support in the aftermath of the harm suffered:

> “I needed a nurse that could be in the house with me. I needed to be able to sleep. I didn’t sleep for months, if not years, because Samantha was such high risk to even aspirating on her saliva. She had an NG tube, even if we fed her that during the night, of aspiration on that. Seizures, you name it. So I needed nursing care in the very early days. I needed that at home.” [Participant 15]

It was clear that practical support was particularly important in cases involving neonatal injuries, where timely intervention is key.

> “while we have funds now for Samantha going forward, which is great, I needed them when Samantha was born. I needed the funds when Samantha was born. I needed the medical support and I also needed to be able to go abroad and to get therapeutic intervention for Samantha when she was small. Like, nought to three is the highest development stage of any child. It’s when the neuroplasticity is greatest. That’s when the money needs to be going into children.” [Participant 15]

Participant 15 alluded to her concern that children whose families who were not in a position to pay privately for assessments and supports before they received financial compensation from the state, which may take many years, would not reach their potential;

> Oh, yes. It’s just… it’s very hard when I look, because I know Samantha is one child… children, who can’t afford to go privately in Ireland and can’t afford to go abroad. And these children are left not achieving their full potential in their lives, and that’s a very difficult thing to have happen. [Participant 15]

Some participants also spoke of the impact of the adverse event on their ability to work and a recognition of the financial implications of this was important.

> Yes, I would have loved if the hospital outside had called and said to me, “Look we are so sorry, we made the biggest blunder, we are going to facilitate your wages while you are out of work and when you go back to work and if any care that you need after that then we will sort that”. [Participant 10]

##### Abandonment

Participants’ frustration with communication received and the lack of engagement and support culminated in a pervasive sense of abandonment. Participant 5 encapsulated this sentiment, emphasising the protracted nature of communication from the hospital as a sign of total abandonment.

> “No engagement. No engagement. If anything, it was the opposite. It took them [the hospital], that letter that I wrote right, it took two months for them to reply to that letter, right. So, we were totally abandoned. Absolutely abandoned…They were covering up a big, a big…you know, they sent me home with the remains of my husband like…and hard luck.” [Participant 5].

Another participant, Participant 11, formed the conclusion that the lack of communication and engagement experienced from the hospital amounted to disrespect and abandonment;

> “And I said, you know, I can’t understand understand why essentially my letter was, I can’t understand why you haven’t contacted me?” which led the participant to conclude that “they disrespected me so much at every turn” [Participant 11].

The lack of communication from her own medical team was also of concern to this participant; *“I have yet to get a call from my obstetrician to see either how I am or how my son is”*. This was particularly conspicuous given that they had engaged with the media on the topic of birth injuries, suggesting that *“people are shopping for evidence for their catastrophic injury cases against maternity hospitals and I’m just like, What?” [Participant 11].”*

The lack of engagement and indifference from the hospital was confirmation of abandonment for Participants 10 and 12

> “ But for them [the hospital], the door closed. They didn’t care, and it was like, yes, c’est la vie. Mistakes happen.” [Participant 10]

> “Like, they could have said, look, hands up, we messed up. This is something that shouldn’t have happened. And we’re going to try and monitor your child now and see if they are disabled, and if they are, we’re going to put them into services and we’re going to help this child get through services. But what happened was we were literally put outside the hospital with the baby, sent home and that was it” [Participant 12]

Some participants acknowledged the impact of the patient safety event on the healthcare professional(s), recognising that healthcare professionals were also often abandoned in the aftermath of patient safety cases. Participant 1 characterised the situation as *“shared abandonment”,* expressing concern not only for themselves but also for a younger doctor involved in her son’s case. In recounting the experience of meeting a Registrar who had been involved in her son’s case, she highlighted how she;

> “turned to my husband and I said, Oh my God, they’ve abandoned us and they’ve abandoned him too. Where did he go? Did he have anyone to talk to? Are they minding him? … It’s almost like there’s a sense that if you’re involved… in such an incident, there was a level of contagion attached to it, and I don’t approve that either.” [Participant 1]

Participant 3 echoed this sentiment and described feeling a similar sense of empathy upon reflection, that the healthcare professionals had also been neglected and left unsupported.

> “Now I can just say as well. I’m being selfish just talking about me and the patients, but like there was zero support for the medical professionals that were in the room with me that time. I left that room hating them. Yeah, over time I’ve come to feel sorry for them. Yeah, because I’ve learned their journey in this as well, which was absolutely thrown under the bus and forgotten about. No support for them which was all wrong.” [Participant 3]

#### Theme 2: The pathway from adverse event to litigation

##### Summary of Theme

This theme delves into the complex journey to litigation following a patient safety incident. Among the pivotal components of this pathway, there is typically an initial pursuit of answers, as individuals seek clarity, accountability and apology in the aftermath of adverse events. The missed opportunities for resolution are discussed as participants identify a range of motivating factors for their decision to pursue litigation.

##### The pursuit of answers

Acknowledgement, truthfulness and timeliness of communication are key components of Open Disclosure. Many participants described their experiences in seeking explanations, both pre- and post-implementation of Open Disclosure in the HSE, as protracted, opaque, unsettling and unsatisfactory.

> “They clearly didn’t know what had happened and if someone had the openness and the decency to say, we have a sense that this shouldn’t have happened and we will investigate it, we will get to the bottom of it and we’ll keep in touch with you about it. And that was all I would have wanted. Because that would have shown a willingness to learn and prevent recurrence” [Participant 1]

Participant 3 in recounting the disclosure of a patient safety incident involving CervicalCheck highlighted the lack of answers provided to their questions

> “when we asked the question, was there mistakes made here? And of course, the consultant could not answer. And I’m sure he was up to his eyeballs and legal advice on how not to answer questions like that.” [Participant 3]

The lack of information and the desire to understand the circumstances surrounding the incident resulted in participants undertaking critical reviews of the patient case. Participant 5, following the death of her husband, described how she and her daughter;

> “had plotted all the little tic tape pieces of information that tell you, you know how the patient, like actually we did in advance of it ever happening, well it probably did happen in hospitals but not to the same extent as we did it. We did a kind of a nearly like a forensic review of Tom’s care of the deteriorating patient, kind of thing. You know, what point did things go, what point did things deteriorate to a tipping point where this was like” [Participant 5]

Participant 11 reflected a similar challenging process of understanding what had happened her child, reflecting a determination to gain clarity and insight.

> “And I kept piecing these pieces of information together… You’re piecing this together, I’m looking it up. I’m googling .. I’m reading medical journals trying to work out what has happened and what the implications are.” [Participant 11]

Participant 15 described feelings of suspicion and frustration following a meeting held with her consultant following the occurrence of a birth injury, necessitating a proactive approach by the participant to seek further information and clarity, which were not forthcoming.

> “So we had looked at that stage for a meeting with the obstetrician. And it was probably maybe a week or so later by the time we had the meeting with the obstetrician, and we were kind of basically told that it was just one of these things that happened, basically. That nothing really had gone wrong. So we requested the CTG and we knew at that stage that the CTG was kind of being around the 100, the 105 mark and there was something suspicious with this… And I suppose the meeting… we got very little, I suppose, answers at that meeting, to be very frank about it.” [Participant 15]

Several participants also described how their journey to considering litigation was prompted by the informal conversations with healthcare professionals which prompted internal reflections on potential system failures or lapses in care.

Participant 2 described how in the aftermath of their child’s death

> “there were others in the hospital who also said that it didn’t sound right and there were people who visited us as well in our house, who basically expressed their concerns, but were not able to.. felt just too intimidated to go on record and speak up” [Participant 2]

Similar accounts were provided by Participants 13 and 15 in relation to the patient safety incidents involving a birth injury:

> “I met Dr A who I could kiss the ground she walks on. She saved my baby’s life. And she basically pulled my Mum aside and basically said, “You’ve got to look into this. This is not right” [Participant 13]

> “Samantha was in the neonatal and, you know, there was, you know, rumblings upstairs, I suppose, comments being passed by staff of, like, these things just don’t happen, what went wrong. You know, that kind of way, I suppose, unofficially and informally. But there was definitely rumblings upstairs.” [Participant 15]

##### Reasons for pursuing litigation

The decision to pursue litigation following a patient safety event is influenced by various factors, as outlined in the literature [Tumelty, 2023]. These factors can be categorised into those with realistic legal aims, such as seeking financial compensation or a day in court, and those with extra-legal aims, including obtaining an explanation, an apology, and preventing recurrence.

However, regardless of their motivations, many participants expressed reluctance towards litigation, emphasising that it was not a decision they took lightly. Several participants, like Participants 1, 4 and 7, expressed an unfamiliarity with the legal system and strong aversion to pursuing legal action.

> “And we’re a family that would would not have wanted to go down that route in 100 years. Our confidence in any hope of ascertaining the truth through honest dialogue was shattered and that is why we undertook litigation” [Participant 1]

> I’ve never taken anyone to Court before. I’ve never been involved in trouble with the law or never anything, you know, so I would know no side of that – nor either do I want to. [Participant 4]

> “We still weren’t going the legal route. We still had no interest in it at all because it upset us to think about it, that… plus all the extra stress and everything.” [Participant 7]

##### ‘Realistic’ Legal Aims

Drawing on Tumelty (2023), we understand ‘realistic’ legal aims as aims that can be delivered by civil litigation. The most significant of these is financial compensation which serves a practical purpose in addressing the additional expense or lost income or opportunities caused by the adverse incident. As we will see, compensation was recognised as important by some participants while for some others it was less important and even served as a disincentive to litigation. The other ‘realistic’ legal aim which is sometimes identified is the desire for a ‘day in court’ which we understand to mean the performance of an acknowledgement of harms suffered in the formal setting of the court room.

##### Financial Compensation

The necessity of financial compensation emerged as a critical consideration for some participants. In cases involving infants and children, the need for ongoing treatments and supports were essential for their well-being and future prospects. Participant 13 emphasized the need for compensation to cover potential therapies and supports for their child, whose future health outcomes were uncertain. Similarly, Participant 14 highlighted the financial burden associated with their child’s medical needs, including expensive treatments. For these participants, pursuing financial compensation was driven by the imperative to secure resources for their child’s future care and support, underscoring the pragmatic considerations that informed their decision to pursue litigation.

> “do I want this to end, or do I…do I fight it for Penny and like I didn’t, at this stage I obviously didn’t know what Penny’s future looked like, and I was thinking if we need money for therapies and stuff,” [Participant 13]

> “So we would have had to pay for the immunotherapy, which is about 40 grand a shot. So from that point of view, that was our driving force because our… our health insurance didn’t cover it either…But no, I think, like, we would have always taken a case … for his future, we were saying he needs this. So I think for his future we were going to do it” [Participant 14]

The need for financial compensation was also highlighted by participants in reference to loss of earnings and the financial burden they had been placed under following their adverse event. Participant 6 described the consequences in her case.

> I said right, this is the rest of my life. They’ve taken my right to work away, future loss of earnings, future… I mean, I wanted to go to college one time, you know. I couldn’t go.

##### Seeking Justice through Financial Awards

Some participants provided perspectives on the role of monetary compensation in seeking justice following patient safety incidents. Participant 13 saw that financial compensation could have potential in holding the healthcare system accountable, emphasizing the need for someone within the system to answer for the monetary damages incurred. Participant 8 expressed how they sought justice through compensation but were sceptical of its real impact, highlighting the limitations of financial penalties on a well-insured healthcare system in eliciting meaningful change or accountability, a view also shared by Participant 9.

> “It being someone, for instance, that money…someone has to answer for that money, do you know, whether they do or not, I don’t know…but it’s…I just felt like that’s the only way I could have got to them. I just wanted to get the answers. That’s all I wanted. I never wanted…you know, a big media thing where they go, “Well we’re really sorry” [Participant 13]

> “my barrister came back and said, “They’re not going to apologise but we’ll make them hurt and we get as much money out of them as possible”…It’s massively flawed, massively flawed because the other thing is that money isn’t going to hurt them either because they’re insured up to the wazoo, you know [Laughs]. Their premiums might go up a little bit but, you know, it’s not going to hurt them. [Participant 8]

> “let’s face it, the law is not about justice, the law is about damage limitation and money… You’re not going to get justice. You’re going to get a payment. That’s it. It’s as cold as that, buts that’s the way the law is.” [Participant 9]

##### Financial Settlement as an unsatisfactory outcome of litigation

As has been recognised by the Supreme Court, ‘an award of damages is an imperfect mode of compensation’ (*Kearney v McQuillan* [2012] IESC 43). The dissatisfaction with monetary compensation as an outcome of litigation was a recurring theme among participants, highlighting the limitations of financial remedies in addressing the emotional and psychological toll of patient safety incidents.

Participant 1 discussed her reluctance to pursue financial damages which underscores a broader sentiment shared by others, emphasising a preference for non-monetary resolutions that focus on accountability and acknowledgment of wrongdoing. Participant 2 made the decision not to proceed with litigation reflecting a similar sentiment, as the prospect of a financial settlement was deemed undesirable. Similarly, Participant 5 prioritised the provision of answers over monetary gain highlighting the inadequacy of financial compensation in addressing the profound impact of patient safety incidents. For Participant 7, the receipt of money as compensation for the loss of their children was distressing, highlighting the profound grief and sense of loss that cannot be assuaged by monetary means.

> “And so I remember the day the solicitor said “And we’ll have to make, we’ll have to claim a monetary sum”, and I said, “but we’re not interested in money, I told you that”, but he said “you have to make a claim. That’s part of how it goes” [Participant 1]

> “we’d end up with a financial settlement..If anything, that was the one thing we did not want.” [Participant 2]

> Like why would I want more money. I had no more need for money. I had enough money, you know. I just wanted someone to answer the questions [Participant 5]

> “we had a big problem with… with receiving money. Oh god, it’s going to set me off again. We just found that part extremely hard because… oh, sorry. No amount of money was going to bring our girls back” [Participant 7]

These accounts collectively underscore the complex and multifaceted nature of seeking justice and resolution in the aftermath of patient safety incidents, where monetary compensation often falls short of addressing the deeper emotional and psychological needs of affected individuals, indeed in some cases, causes further distress.

##### Day in Court

Contrary to some findings in the literature suggesting that a desire for a "day in court" is an aim of some plaintiffs seeking justice, most participants in this research did not express this sentiment. Participant 10 adamantly stated their reluctance to pursue this route, expressing a clear aversion to the courtroom setting, a view also held by Participant 14. Similarly, Participant 11 emphasized the reluctance of parents, whose children have experienced catastrophic birth injuries, to engage in litigation, highlighting the emotional burden and desire to avoid reliving traumatic experiences. However, Participant 13 expressed regret over not having the opportunity for a day in court, seeing it as a means to seek justice and hold responsible parties accountable in a public forum. This sentiment reflects a nuanced perspective within the participant cohort, with some recognizing the potential impact of a courtroom setting in achieving broader accountability and awareness of patient safety issues.

> “I didn’t want to go to the Courts. No. It wasn’t going to be me going to go up on the stand and things like that, no” [Participant 10]

> “We would have settled definitely, because I didn’t relish the thought of going into the High Court and standing up there for an hour, you know, talking, like. I didn’t. You know, I wasn’t looking forward to that at all, you know, as you can imagine, you know. So yes, I would if I could have avoided it, or we could have avoided it, yes” [Participant 14]

> “Like no parent with a child with catastrophic… This is what nobody seems to understand. None of them want to be in the courts. None of them want to be in a litigated process. None of them want to have to relive it.” [Participant 11]

> “I…there’s days where I regret that decision, that I wanted my day in Court and I wanted them to answer. I kind of, not that I wanted the media involved, but I wanted it to be publicised that they had done something majorly wrong, and I felt maybe that I was a voice for maybe some other women to come forward and, you know, to not be as afraid” [Participant 13]

##### Extra-legal aims

Participants also identified a range of ‘extra-legal aims’, aims which the legal system is either less able or entirely unable to deliver.

##### Explanation and Seeking Truth

Seeking an explanation and uncovering the truth emerged as a primary driver for many participants considering litigation following a patient safety incident. Participant 13 was clear that had she received answers and satisfactory follow-up for her child, litigation would not have been necessary. Participant 3 expressed a sense of necessity in pursuing legal action to obtain answers regarding his wife’s death, emphasising the failure of the litigation process to adequately address the need for transparency and truth-seeking. Similarly, Participant 5 emphasized the importance of receiving answers and viewed the legal system as a means to achieve this goal. Participant 7 explored the factors her family balanced when deciding to pursue litigation in their search for answers. These participants articulated a desire for accountability and acknowledgment of system failures, underscoring the significance of seeking explanations through the legal process

> “had I got the answers and some sort of explanation, or some sort of treatment plan…and not be taunted, and not be…I would have never gone down the legal route.” [Participant 13]

> “the majority of time you just want to know what happened. But of course, your only way of getting it… When we asked the HSE, when we ask the consultant, like they just said we don’t, we don’t know..I needed to get to the truth. And I guess probably another part of the whole failure of the whole litigation process is where you are forced down the legal route.” [Participant 3]

> “I just wanted someone to answer the questions. I wanted someone to be, to say well, our system was flawed in that particular day. Like I found out so much in dribs and drabs over the years.“[Participant 5]

> “trying to find out the truth. We had… that’s what we had to debate about, you know. Do we lose the house with our two children that we still have or, you know, do we find out the truth and, you know, we had to weigh up everything. And, like, how could we tell our kids now later on that we didn’t fight enough and didn’t find out the truth about their two sisters, you know?” [Participant 7]

##### Apology

A key component of Open Disclosure is the apology to the affected person(s) following a patient safety incident. This study found that the absence of an apology for the occurrence of the event and the treatment of patients and families thereafter was a significant driver in pursuing litigation. Participant 6 emphasized that obtaining an apology was their primary goal from the outset while Participant 2 underscored the value of receiving an apology for the treatment they had endured. Both participants conveyed a clear desire for an apology as a crucial step towards resolution and healing in the aftermath of adverse events.

> “I wanted an apology from day one. That’s what I said, in all my assessments, I want an apology.” [Participant 6]

> “We wanted an apology. That was really important for the way we have been treated. Uh for gas lighting us and you know, like we didn’t really care how deep the apology was. We just wanted an apology of some sort from the hospital” [Participant 2]

##### Preventing Recurrence

Some participants demonstrated a strong sense of responsibility towards others and a desire to prevent the recurrence of similar incidents, viewing litigation as a potential avenue to achieve this goal. Participant 11 expressed deep concern about the possibility of similar events happening to others, emphasizing a personal duty to ensure that such tragedies were avoided in the future. Participant 6 articulated a desire to uncover the truth and implement preventive measures to safeguard against similar occurrences while Participant 13 also expressed a motivation to pursue litigation to promote awareness and change within the healthcare system. Participants conveyed a commitment to advocating for systemic improvements and ensuring accountability to mitigate the risk of similar patient safety incidents in the future.

> “ I’m worried that this is gonna happen to somebody else, and every time for years, I see ‘dead mother, dead baby, not listened to’. It’s always because the mother wasn’t listened to… . I am responsible. I’ve got to make sure this doesn’t happen to somebody else.” [Participant 11]

> “All we wanted to do was find out the truth. And, you know, make sure that, you know, what happened was never going to happen again, or at least, you know, that precautions were put in place and everything that needed to be done was to the highest…to make sure that cases like ours weren’t going to happen again. That was the only interest we had.” [Participant 7]

> “And I just didn’t want this to happen to someone else… And that’s what I’m all about. You know, trying to pay it forward a bit, because I feel like I’ve got a miracle at home.“ [Participant 13]

##### Accountability

Participants emphasized the importance of accountability and ownership in relation to their patient safety cases, expressing a desire for acknowledgment and acceptance of responsibility from the healthcare professionals and institutions involved. Participant 10 articulated her motivation for pursuing litigation, highlighting the significance of the consultant acknowledging their mistake, offering a genuine apology, and committing to preventing similar errors in the future. Similarly, Participant 2 underscored the need for the hospital and consultant to acknowledge and accept what had occurred, emphasizing a desire for transparency and accountability in the aftermath of the incident. Both participants sought validation of their experiences and a commitment to improvement from the healthcare providers involved, reflecting a broader aspiration for accountability and learning within the healthcare system.

> “Yes, so that was my thing of taking the case initially was, for her [the consultant] to stand up and say, “Yes, I made a fault, I made a mistake, this is why I made it and I’m so sorry for it, it will never happen again” [Participant 7]

> “We just wanted to find we just wanted to understand what happened and we just wanted the hospital and the consultant to accept what had happened.” [Participant 2]

##### Advocacy for Women’s Health Equity

Several participants in the study voiced strong concerns about the treatment of women within the Irish healthcare system, highlighting systemic issues and advocating for significant change to prioritise women’s health needs. They expressed frustration with what they perceived as a patriarchal system that often dismisses or diminishes women’s concerns, particularly in the context of childbirth and women’s reproductive health. Participants emphasized the need for medical professionals and healthcare institutions to provide empathy, compassion, and dignified care to women, regardless of their background or circumstances. For several women, this was another factor that motivated them to pursue litigation.

> “There’s a beautiful Irish word, “flathulach”, I remember my GP in Dublin saying that men doctors, particularly, are just a little bit flathulach with women’s health - and I think the Irish government and system just doesn’t take our health as seriously as, I mean, you never hear of a testicular cancer scandal or a prostate cancer scandal, you know, it’s always just, it’s always women, you know, let’s saw people’s pubic bones in half to get the babies out, you know, sure, it doesn’t matter if they’re disabled for the rest of their lives, just whip out their uteruses, you know, second class citizens when it comes to health.” [Participant 8]

> “the point of the maternity hospital is to care for women. The point of the maternity hospital is to care for women, all women and and you know, if you’re a traveller woman in Ireland, if you’re me, if you’re… like, you should just be treated with dignity and care and kindness, you know, it’s not… Like there’s something that remains very patriarchal, even by the women. Do you know, there‘s something very dismissive and patronizing instead of the woman being that the centre of things. You see, I think once the baby is born, it’s the baby… instead of the woman being and her care and her well-being.” [Participant 11]

> “I’d change how women are… I would change how women are educated about when medical negligence occurs. Like, we have women in this country who when they have babies, they wee everywhere. That’s not normal. Like, we have women who when they sneeze and cough, they wee. That’s not normal. That’s not supposed to happen. But our medical profession is telling us that’s what happens when you have a baby. Or what happens if you have a painful period and you’re 16? They tell you when you have a baby, you’ll be fine. You mightn’t have another baby for 20 years. So I think the way we treat women in this country medically needs to be changed. I would change how the medical profession and… treat women. I would change how we refer to women in documents, particularly mothers. Particularly first-time mothers. Any mother.” [Participant 12]

##### Deficiencies of the litigation route in addressing extra-legal aims

Several participants recognized the limitations of the litigation route in addressing their extra-legal aims. Participant 13 acknowledged the unlikelihood of receiving an apology through the legal process, prompting them to accept the absence of this form of redress.

> “I knew we were never going to get an apology, first of all, and that’s what my lawyer said to me, “You’re never going to get that”, so…you’re got to just deal with that.” [Participant 13]

Participant 2 expressed their reluctance to engage in litigation, as despite the strong possibility of a settlement in their case, they recognised that it would not necessarily result in the hospital admitting liability or offering an apology, which was a crucial aspect of their pursuit for closure.

> “we didn’t make a claim like one thing was we didn’t want it to be about money and…they said there’s a chance that the hospital would settle out of court and we still… they still wouldn’t admit any liability. ….So there was no benefit to us” [Participant 2]

Some participants who opted for litigation emphasized the importance of achieving extra- legal aims as part of their settlements. Participant 1 recounted the process of settling her son’s case, highlighting that while financial compensation was not their primary concern, the admission of liability and acknowledging the negligence that led to the incident was crucial. This acknowledgment served to absolve her son, as his condition manifested in behavioural symptoms, and underscored the importance of holding the responsible parties accountable. Similarly, Participant 6 prioritised receiving an apology during the settlement negotiations, emphasizing that monetary compensation was secondary to obtaining acknowledgment and closure.

> “But anyway, that all went back and forth, and again, as I say, we weren’t interested in the money, but eventually they did concede liability… I felt that the admission of liability vindicated Kevin in that his unacceptable behaviour etc was uncharacteristic and was driven by his undiagnosed condition” [Participant 1]

> “No, I won’t be saying any numbers until I get an apology”. So, they went off and they looked for an apology. They came back and said there’s a strong possibility you will get it and I said, “Oh my God”, ok, this is all I wanted, I didn’t want anything else.” [Participant 6]

##### Financial Risks of Litigation

Participants grappled with the significant financial risks associated with pursuing litigation, highlighting the profound impact it could have on their lives and families. Participant 1 and 7 candidly described the daunting prospect of potentially losing the family home if they were unsuccessful in their case, underscoring the immense stress and difficult decisions involved. Despite the financial strain, their determination to seek answers and justice compelled them to proceed, recognising the high stakes involved. In contrast, Participant 5 opted not to pursue their case due to the overwhelming financial burden and the fear of losing their livelihood. The prospect of further financial losses compounded the already devastating emotional toll of their loss, leading to a difficult decision to forgo legal action despite feeling aggrieved.

> “Because you know both in areas of finance and resources, and the plaintiff takes well, we took huge risks. We didn’t know whether we’d still have a roof over our heads at the end of the day, but I remember walking in this door and saying to my husband, do these bricks, do these bricks and mortar mean more to me than Kevin? No, they don’t And there was a very strong resolve there from day one.” [Participant 1]

> “We had to make the decision that we could, at the end of it if we didn’t win our cases, we could lose our house. And we already… we had two other kids as well. So we had a lot of thinking to do there… Yes, it was very stressful… But our fight to find out… we just needed to know. And if we were going to lose everything else, we needed to know, and we needed to fight for it” [Participant 7]

> “So, that’s why I didn’t proceed. It wasn’t that I didn’t feel aggrieved. And I was like, Tom’s dead, I don’t, if I mind my farm…, I was afraid I’d lose the farm as well… So, I just started to see myself haemorrhaging money and what was I, it was €2,000 to bring, to get the senior counsel at the inquest so it was all you know… And I thought I’m never going to get Tom back. If I lose his farm on top of him, I’m just going to slit my throat.” [Participant 5]

##### Missed opportunities for mediation and resolution

The narratives presented shed light on missed opportunities for mediation and resolution within the healthcare system, emphasizing the importance of fostering a culture of openness and accountability from the very earliest time possible following the patient safety incident. Participant 2 underscored the necessity of creating an environment where healthcare professionals feel supported in disclosing safety incidents and acknowledging potential errors. Participant 5 echoed this sentiment, lamenting the adversarial nature of the litigation process and highlighting the potential for open and honest conversations to prevent further harm. Similarly, Participant 7 and Participant 1 expressed the need for Open Disclosure, emphasizing that transparent communication and genuine dialogue could have provided the answers and closure following the death of their children.

> “we need the the environment needs to exist to allow a person to go back and say, look, I could have got it wrong. Here’s what I think might have happened.” [Participant 2]

> “It’s just, no matter what side of the fence you are on this, there’s no winners like. It’s all losers, and if you can find that place maybe where there’s reconciliation where you can sit down and have the chat. If I could only extend that first day when the surgeon was crying for us and I was crying for him, if we could have kept that going for a lot longer, I wouldn’t have gone, I wouldn’t have gone through the court system. I wouldn’t have felt the need.” [Participant 5]

> “Open disclosure, for a start. Because there was none of that in our situation, in any way, shape or form. Just, as I said, open disclosure. Sit down and actually have a conversation with these people and find out what exactly happened. Like, it happened in… you know, if we had known exactly what had happened in Hope’s case, and obviously we would have been extremely angry, really upset, you know, if we were told at the time, look, mistakes were made, you know, Hope should have had a chance, she should have been delivered three to four hours earlier than she was, she should have had a chance. You know, I know for a fact we would have walked away and just… this is what happened. It wouldn’t have even entered our minds to take legal action at all. Because in our minds that’s like, what’s that going to achieve? We’re not going to get Hope back. [Participant 7]

> “They clearly didn’t know what had happened and if someone had the openness and the decency to say, we have a sense that this shouldn’t have happened and we will investigate it, we will get to the bottom of it and we’ll keep in touch with you about it. And that was all I would have wanted – an honourable and decent encounter with individuals and answers to our simple questions – why did he die?” [Participant 1]

These accounts underscore the potential of Open Disclosure in averting the trauma and distress associated with litigation, highlighting the importance of proactive measures to address patient safety concerns and promote healing and reconciliation within healthcare settings. As described by two participants,

> “It shouldn’t be the adversarial thing. You know where it is about blame and so on. And we didn’t want it to be, you know, we wanted it to be about open disclosure and about understanding and empathy and and so on.” [Participant 2]

> “I think that has huge potential for healing on both sides” [Participant 1]

#### Theme 3: Experiences of the Legal System

##### Summary of theme

This theme explores the experiences of participants of the legal system, including their experiences with legal professionals representing the state and their own legal teams. It explores the sense of loss of control experienced by those who took legal action against the state, of being pawns on a chessboard, ancillary to the process. Participants also describe the lengthy nature of the process and the failure of the system to meet their needs. This research demonstrated that the outcomes which plaintiffs are willing to accept are altered by an adversarial legal system (which is centred on financial settlement) and the legal profession (who are indoctrinated in a culture of victory and defeat which is measured in monetary terms). The need for communication with, and education of, those engaging in legal proceedings was highlighted with participants demonstrating a lack of clarity about how their cases were settled and the distinction between settlement talks and mediation. The confusion is perhaps unsurprising given that even when mediation was attempted, the process described by participants did not embrace the full potential of that medium, appearing to be nothing more than settlement talks facilitated by a third party. The toll of participating in assessments as part of the litigation process emerged as a strong theme of this research and will be explored fully in the next section exploring the emotional and mental impact of litigation.

Participants were unified in their view that their experience of seeking justice through the civil litigation system was negative, describing it as triggering (Participant 4), re-traumatising (Participant 4), jarring (Participant 4), insulting (Participant 14), shocking (Participant 14), horrendous (Participant 3, Participant 6), not fit for purpose (Participant 3), horrible (participant 10) and a battle (Participant 13). Several participants identified the system as particularly unsuited to those who have already experienced a significant trauma. Participant 15 considered the experience of taking legal proceedings to be more traumatic than the original adverse event

> “I’d be very honest, like. Obviously, Samantha’s birth was traumatic and took everything out of me, obviously, and to be left with a child and everything. But the legal proceedings, especially the last two years, were actually worse than Samantha’s birth and I say that very honestly.” [Participant 15]

> “the legal process is a horrible process. It’s unfair, it’s not a place for people to be who’ve been damaged.” [Participant 5]

> “The experience, I suppose overall, it’s not a very happy one. It’s not a very good one. You know? I mean you’re shoved into a world where you no nothing about. It’s very jarring. It’s very, I suppose, re-traumatising. It’s very triggering.” [Participant 4]

The experience of Participant 6 highlighted that the trauma can be experienced even in the final stages of the process. Describing her experience in court when her settlement was approved:

> “So, I agreed and…to settle, there and then for that, and then I had to go into the court, meet the Judge and the Defence and I went in, and I will never, ever forget it. I felt dirty, victimised…it was terrible…. He….my own barrister… he said my name, and my husband’s name, because he was co-Plaintiff and he never even looked out at us. There was no acknowledgement, nothing.” [Participant 6]

The strategies adopted by opposing counsel on behalf of the state were commented on by several participants.

> “And then when we went to court, again it was like… it was like… it was insulting. It was like… it was insulting to my intelligence really, you know, the games that were being played. I was thinking, oh my god, you know, like, how were people being let away with it. So there was kind of this, like… skullduggery nearly going on.” [Participant 14]

> “Mr H [Barrister], I just felt he was going to the lowest of the lows, you know, like. He tried everything. He was… it was… it was just shocking, you know. So yes, really shocking.” [Participant 14]

> “They wasted so much…they wasted so much, so much of my life, you know? It was dirty tactics, they were just – they were just trying to reduce their liability, in any way at all possible, and in the end they rounded the whole thing up.” [Participant 9]

> “I wrote to them, my solicitor wrote on Jack’s behalf and I said, and she’s basically said, look, we want to do everything we can to avoid litigation. You know, if there’s a way of resolving this by mediation, … she’s saying my client wants to reduce the cost of the state. The costs, the litigation, everything, have the least adversarial model possible. Can we agree x and y and z. And notwithstanding all of that … they still said, but this might be caused by [X]. It’s just and I mean, I was… I said this in the High Court when his case was being ruled and my barrister was saying like this has been rubbished by all of our experts and they’re still saying this. I was saying it just hurts so much because not only did they not take responsibility for it at the time, not only did they not treat me with any respect, any kindness, any dignity and but to still be making this position” [Participant 11]

##### Experience of engaging with their own legal team

In contrast, participants broadly described very positive experiences with their own legal team.

> “They tried, they worked very hard for me and they were very reassuring all the way through it” [Participant 8]

> “you know my legal team, I think are the most fantastic, amazing people and the most undervalued people and professionals as well. Because they just couldn’t have been more sensitive and more willing to get me while I want with regards to truth.” [Participant 3]

A number of participants reported finding the empathy and understanding they encountered in this sphere to be in contrast with their experience in the healthcare system in the aftermath of the adverse event.

> “fortunately the consultant anaesthetist [expert witness]… we covered his travel and his hotel and that was it, you know, he didn’t charge anything. Uh, the the barrister waived his fees as well, which was unbelievable. Umm, the lawyer waived her fees. So you know, we were incredibly fortunate, but you know, if we had seen that level of empathy and compassion from the the hospital and the medical professionals, then, you know, we wouldn’t be in this position. You know, we would have been able to understand what had happened.” [Participant 2]

> “No, my experience of the hospital and the health services is horrific. But my experience of the legal… And he was actually very personable and he was personally involved with Freddie. Like, he knew us, if that makes sense. So I found him to be very effective. He was lovely. He was a lovely man, I have to be honest. All them were. All the guys were.” [Participant 12]

While their overall experience was positive some participants pointed to a lack of communication with their legal team.

> “Well, my solicitor was brilliant. My solicitor was actually excellent. And they didn’t really bother me at all” [Participant 12]

> “my Solicitor was very good and very thorough, and my legal team were very good and very thorough. But in general, you know, either they don’t have time, or they chose not to kind of give you a project, project update or status updates. So like I had to constantly go and try and find out, where are we, what’s the next step, and we…any time any information was requested from us we had it, you know, within 24/48 hours, but…”[Participant 9]

One participant outlined a very negative experience with her legal team.

> “But it shouldn’t have been the battle we had, and I suppose we also had a lot of disagreements with the prognosis and the diagnosis that our own solicitors sometimes were putting on Samantha… It came down to financial gain for them too sometimes, as opposed to the truth about Samantha, because she had done so well…. And I suppose our legal team were looking to kind of more paint a diagnosis of cerebral palsy… They did get an expert report of that she had subtle… I suppose, subtle signs and flags of cerebral palsy. So once the word cerebral palsy was on the report, it was enough, even though it was subtle… it’s very difficult when one of your biggest disagreements and so forth is with your own legal team.” [Participant 15]

Participant 15 noted that it was in her legal team’s interest to settle the case out of court rather than allowing her to take the stand.

> “I suppose… our own team didn’t want this to go to trial either because they didn’t want me to go on a stand when they knew that I didn’t agree with the diagnosis that they’d gotten from their expert witnesses. So there was always that debate of trying to keep me quiet, even from my own legal team, because I would have been quite outspoken to them and I would have spoken to them about my grievances with this. So there was always a push that mediation would happen.” [Participant 15]

She explained that she felt bound by her contract with her legal team and forced to acquiesce to their strategy.

> “I couldn’t get out… of the contracts. That is why. If I tried to get out, I had to pay their fees, so we’re looking at a six-figure sum that I’d have to pay them to get out of the contract.” [Participant 15]

##### Loss of Control During the Legal Process

Several participants, despite maintaining relatively positive connections with their legal teams, expressed a sense of losing control once the legal proceedings commenced, instead they were now part of an ongoing and inevitable process. Participant 6 describes how they *’just went into a bubble of figures. It just turned into this… like a horse mart. Like an auction online for a racehorse.’* For this participant, the system took over and *‘the person gets lost, the real person gets lost in it.’*

For Participant 13, it felt like their child had become lost:

> ‘*when they talk about Penny, they talk about it as if she’s a number and I’m like, “She’s right there, she’s my baby”, you know.*’Participant 1 described becoming sidelined in “a *joust between the experts*”. It “*came down to the cleverness of these people and the experience of these people and really with no great input from ourselves.*” The highly emotional nature of the process was highlighted;

> “it was this sense of that it’s out of your hands, and because he was so precious to us, and because the whole incident was so appalling, I know I for one, didn’t like this feeling of being sidelined because part of our healing was our doing our bit of the job and when we hit this point, it looked like we had no role to play. And it was that bit again that we were kind of at the mercy of yet another system.” [Participant 1]

##### Power Dynamics

The challenges created by the structural imbalances inherent in litigation by an individual against a hospital and/or medical professionals were highlighted in this research, which Participant 1 described as a *“David and Goliath experience until the 11th hour”.* Participant 3 echoed this sentiment when they described feeling that *“It’s you against the HSE or it’s you against the system.”*

Participant 1 described a legal system which favours the defendant *“both in areas of finance and resources”* and the financial risk taken by participants is explored further in theme 2. However, Participant 1 recognised that some are not in a position to take a financial risk of this nature and was deeply affected by the injustice caused by the exclusion of those without the requisite resources from pursing legal action as a means of achieving justice;

> “So we wrote the cheque for £2000 and I remember when we came out of his office and I stood with my back against the wall and I was crying and my husband said to me, why are you crying? Sure we’re doing something about now. I’m crying for the mother who doesn’t have £2000. Desperate because there are people who are totally disempowered by finance, by other things, that’s where I get back to, the defendant has all these, the resources and so forth, where you were really at the mercy of the system that you know very little about.” [Participant 1]

Participants also described feeling intimidated by highly educated professionals. Participant 13 described the process as daunting and Participant 6 described feeling vulnerable:

> “it was very daunting, because you’re going to someone who is highly educated and you’re trying to fight for your child’s, you know, what’s happened to you and your child. And you kind of feel like you’re a complainer, you know that kind of way, and I’m not! I just want better for people, you know.”

> "I think of me as that distressed person coming in and just going, please, someone just help me here or guide me. We are not a family that has any sort of…I wouldn’t say we’re not knowledgeable or intelligent, we’re just not professional, or we don’t have professions…you know, we needed help, and we needed some sort of guidance” [Participant 13]

> “because you are so vulnerable. You are so open, and you’ve got these highly intellectual people that are there, and you do feel small. I felt so small really. Even though I felt so big at the same time, because I….I was powerful because I had my apology, but at the same time I felt such a small little person in this conference room because of the legal world and all these…you know.” [Participant 6]

> Like, I’m not…I wouldn’t say I’m that academic, but I’m not stupid either, like, I just wanted to get some sort of common knowledge about it. [Participant 13]

One participant spoke about how her medical knowledge gave her an advantage in the legal system which is not available to many litigants;

> “ just felt like, you know, I was in their playground but they were talking about stuff that I know about, so… I was confident in my language and I just feel like had it been, like, my sister, who has no medical knowledge, I just think, oh my god, you know, like she would have been bamboozled and you would have been that much more nervous. You know, like, obviously, as I say, I was nervous because I was in the High Court, but I wasn’t nervous about speaking because, you know, I knew what I was talking about. But for somebody, a lay person to stand up there and try to explain something that they’re not maybe a hundred percent sure on what they’re talking about must be really difficult.” [Participant 14]

##### Failure of civil litigation system to meet plaintiff aims/ Alteration of participant’s aims during the process

Several participants pointed to the fact that the civil litigation process, focused as it is on financial compensation, did not meet their aims.

> “It was like everything was all about money, and that was…. And I’m going…what do you mean, but what’s going to be the outcome of it? … So then you telling me, “Oh this is the price that they’ve decided that’s been put on you”. Everything after that they didn’t care. They didn’t care what happened to you, they didn’t care the reasoning behind it, how it happened, or what was going to be put in place going forward for our kids…There was nothing of that. It was a thing of, here’s some money, go away and shut up now.” [Participant 10]

Participants described having to withstand pressure from a legal system where success is measured in monetary terms and legal professionals are indoctrinated in this culture, focusing on the value of the settlement above other aims;

> “He said, “they won’t give the liability”. Well, I said, “we’re going into court tomorrow, if they don’t”. But he said, “I think I could get more”. But I said “we’re not looking for more, but I could see in his face that he wanted to do better. Because any settlement was being donated to two charities. I said “Well for every 250 punts extra there will be another piglet going out to Malawi with Bothar”. [Participant 1]

They described their legal team guiding them through the process, ensuring that they received financial compensation, irrespective of their other aims.

> “I said, “I want an apology”. Right? And it was on a Zoom, and they were looking at me, and my own solicitor rang me afterwards and they said, “Jane, you have to mention the quantum” and like they gave me a figure… And I was like, “but I can’t, I can’t say figures. I can’t because it’s not…” but they said, “Jane, this is the, this is the whole bottom line of it”. And when she said that, I thought, “oh my God, this is…”, I knew it was the bottom line but just to admit it to myself because all I wanted was an apology.” [Participant 6]

> “Word came through then; I got my apology, and they said I can’t go any further until I’m happy with the wording of it. Now I have to say, it was a very genuine normal written, no bullshit apology. Hand-written from him. And that’s all I wanted. Nothing else, nothing, I just wanted an apology and I wanted to find out what happened. But of course, being the legal system, you…when…you know, you have to go further. .” [Participant 6]

> “the barrister that was there with us, he was saying, like, how much are you hoping to get, and myself and my husband… because our solicitor had even warned us before we went in, don’t be saying, oh, you don’t care, you don’t mind, because she knew what we were like. Because she knew that, you know, we had a big problem with… with receiving money.” [Participant 7]

##### Prolonged Nature of the Process

Some participants resolved their cases in a relatively short time frame (Participant 8 settled within 25 months, Participant 14 within 33 months), often due to the perseverance of their legal team.

> “It was short. The solicitor was like a dog with a bone. She was in the High Court every week before the case. So… oh god… I’m going to… Ciarán was diagnosed 2020. We were in the court in 2023. We were in court in 2023… So it was quick.” [Participant 14]

However, for most participants who took legal action, it took significantly longer to resolve *[For example, Participant 1, 5 years; Participant 3, 4 ½ years; Participant 9, 5 ½ years; Participant 13, “nearly four, five years”; Participant 15, 8 years to first stage]*.

It was clear from this research that participants found the prolonged nature of the process to reach a settlement difficult.

> “it was nearly four, five years…of a legal, when I say legal battle, I mean battle.” [Participant 13]

> “the process, it took 4 1/2 years when we knew the truth from about a year in. Once we had examined everything. And we’re like, we didn’t need to waste that other 3 1/2 years you know. It doesn’t make sense. And I know that everyone’s busy but look this is the problem.” [Participant 3]

Some participants expressed the view that opposing counsel delayed the process, despite their desire to reach a quick settlement.

> “But as far as the State’s Claim, it was just disgraceful the way they dealt with it. Then down to, like, my solicitor was pushing for this to go through… you know, this case to go through. And they kind of did everything, I felt, to stall it.” [Participant 14]

> “They just, they tried to delay and they tried to break me, they tried to break my spirit.” [Participant 13]

> “I found even when they [HSE] were filing, like, and when they had to file the Notice of Summaries and all that kind of stuff, they left it to the very last minute. They were always, they were always over…they were never on time, and that just…to me, I’m like, you’re still disrespecting, you know? Instead of actually facing it, you’re not facing it, that’s what I always felt.” [Participant 13]

> “It took five and a half years to do a six-month job. That’s what I think of it. Because the Defendants did everything to delay, obfuscate, and just in the end, after four years they…after four years of having been asked they finally, they finally under a Motion through the Court, they finally submitted their Defence in which they said that their sister company was liable. So they sat on that for four years.” [Participant 9]

There was also a sense amongst some participants that professionals involved in litigation may have been incentivised to prolong proceedings;

> “Plus they cost thousands as well, on top of, you know, on top of the trauma. So to us, you wonder is it another money racket then as well on top of…” [Participant 7]

> “ I didn’t understand how long it was going to take to get it closed off. But, there’s absolutely no need – five and a half years – you know? Somebody is making a whole pile of money out of this, and it’s not me.” [Participant 9]

Participants also explored the impact of the delays on them and their family members and this will be discussed in more detail under theme 4.

##### Litigation strategy when liability admitted

The narratives presented highlight a dissatisfaction with the litigation strategy adopted by the state, particularly in cases where liability has been admitted. Participants felt this was a waste of state resources and added significantly to the burden experienced by those impacted by an adverse event.

> “It should have been a case of, you know, “Yes, we’ve screwed up. Here’s your letter. We’re really sorry and here’s what we’re going to give you for it” [Participant 8].

> I just think when something is such clear cut. Two people had said yes, we’ve made a mistake, you know… Court, I just don’t think we should have been there. Just plain and simple, I just think it was an awful waste of people’s time, it was an awful waste of public money. And it was just a bit of a joke, like.. Yes, my big bugbear was I just thought it was an awful waste of money, us being in court for three days. [Participant 14]

> “do you know, and having to go up on the stand and go through every minor detail of each stage that you’ve been through and are going to rip shreds off you. I think that’s the most demeaning thing to do to any, any human being. Especially when they know they’ve done wrong… Just pay it out. Just give it to them. Don’t be so disrespectful and… dragging them through the Courts and having to relive every minute that they’ve been through this. It’s horrible” [Participant 10]

> “I suppose, do you know what, overall is, when you’re told that medical negligence has been caused to you… Why should you have to go through barbed wire. Barbed wire and get torn asunder to prove what, you know, to prove what was done wrong to you. Put your hand up. Put your hand up and say right, this is it, but it’s all about money, quantum. That was it, nothing else, all about money. And the person definitely gets lost in it. I got lost in the system. .” [Participant 6]

##### Perception of litigation and litigants

Participants in this study explored the perception of litigants amongst the legal and medical communities and the wider public, pointing to an assumption that litigants are interested in monetary settlements. It was evident that participants felt that litigants were viewed negatively; *“[t]hey may decide to go for litigation and that is their right and you can’t blame them for that and you can’t think less of them for that.”[Participant 1].* Participant 6 described her perception that those involved in her ongoing medical care were focused on the monetary settlement she received:

> “And on the next visit I went, and her nurse and the gynaecologist said to me, “so did you go on a family holiday after the case? Where did you go? Did you go on a cruise?” Oh - my - God. I couldn’t believe it” [Participant 6].

Participant 1 described how she felt that the family’s decision to donate their settlement directly to their chosen charities;

> “made us a kind of, I was going to say interesting, but more of an oddity, because we didn’t fit the perception they had of litigants. It just didn’t fit the profile that they had. Like we’re not, we’re not greedy. We’re not people who are looking to profit like. It didn’t, it just didn’t fit and and I think that that’s the bit that’s kind of confounded them in many ways.”

Even during the litigation process, given its length, some birth injury litigants were made to feel they were fraudulently pursuing money because their child had ‘*done too well, so had her birth really happened’ [Participant 15]*

The stereotyping of birth injury parents as potential litigants ‘out to get the system’ seemed also to hinder open disclosure:

> So from there then we met [Dr Y], and she basically told us that like at two points… I think you should have been sectioned. so I asked him could we get that in *writing, and she was like, “Why?” And I said, oh I just had legal advice that…and from there then it was closed doors, yes. She wished me well and, now she wasn’t rude to me, but she certainly wasn’t helpful. [Participant 13]*

Several participants reported how they felt that the attitude of medical professionals towards them had changed, becoming colder and more distant, because they had taken a legal action. Participant 3 described this feeling as like an ‘Iron Curtain’ coming down:

> “It’s you against the HSE or it’s you against the system. And you know, when you walk into a hospital, the medical professionals are around you and want to do whatever they can to help you. But God forbid something happened and you accuse them of something. They’re no longer your friend.” [Participant 3]

Participant 4 explained that they felt that *‘medical professionals are very, are less trusting of me.’* This participant, who has ongoing health issues, described an encounter with a new medical specialist:

> “[S]he more or less dismissed me, kind of, at everything, when I was asking questions and…like, I never once mentioned about [my legal case], but I know that they went in to get a report then towards the end of my journey, so she would have known that it was from a solicitors anyway … and so I felt that she was immediately dismissive, and got her back up, and you know, and things like that, and it was just, it was not a pleasant experience. Not a pleasant experience.” [Participant 4]

##### Mediation not operating as intended

The experience of participants in this group seems to point towards a system where mediation is not being utilised to its fullest. Participants recognised the potential for mediation to facilitate better communication and produce better outcomes for all involved:

> “I think mediation because people are, you know you’re dealing with broke; you’re beaten, you’re dealing with broken people on both sides… You have to, and a safe place to, a safe place to express your expressions and not, not lose that communication, not break that strand of communication.” [Participant 5]

However, opportunities to engage in mediation with participants who recognised the potential for mediation to fulfil their aims following an adverse event appear to have been lost in some cases.

> “Court, I just don’t think we should have been there. Just plain and simple, I just think it was an awful waste of people’s time, it was an awful waste of public money. And it was just a bit of a joke, like.” [Participant 14]

One participant described attending an arranged mediation, but the opposing side did not attend

> “So the surgeon and the pathologist involved admitted liability, which we thought that would be it. But no. We ended up in the High Court… we went to mediation or we arranged to go to mediation and I went to some… myself and my husband went to some hotel in Dublin. They didn’t turn up for the mediation”. [Participant 14]

While some participants did engage in a mediation process as part of the resolution process, it seems that the mediations as described, are mediations in name rather than spirit. They do not reflect best practice, with little involvement of the litigants, little exploration of alternative outcomes and often taking place many years into a litigation process.

> “we sat in a settlement room and there was a notional mediator but like we didn’t all sit in the same room or anything like that, you know… there was me in my room with my lawyers who then left to go and talk to the mediator. Do you know? Like she might as well not have been there. The focus was interim… care costs, so… No, it was pointless.” [Participant 11]

> “So, it was like mediation. So, I got onto a. Zoom call or whatever, maybe Teams with my barrister and a mediator who had done a lot of the cases. I never saw the other party, at all, okay? ….. And they were just, again, like, very jolly, you know, barristers enjoy this kind of a case because it’s, sort of, easy really…This negotiating, that part, that was very quick, done and dusted within about, like, six hours.” [Participant 8]

> “I don’t know what went on behind closed doors because we were in a room on our own. But some conversation had to happen behind closed doors there even with the mediation, kind of, the HSE were in one room, our legal team were in one room and we were in one room. So while we were supposed to be the centre of it, we were still very much on the outside of it. You know, which was very difficult because, like, our own legal team were in the room for four hours and didn’t come to even speak to us, do you know. So, like, this was about us and about us having mediation and we were very much, I suppose, bystanders and spectators in this.”[Participant 15]

This research has also highlighted the need for clearer communication with plaintiffs about how the case is proceeding, including how the parties to the case are attempting to resolve it. It was evident that many participants were unclear about how their cases were resolved, with several participants describing what appeared to be settlement talks as mediations.

> “basically they two barristers spoke with each other, there was nobody really spoke with me. It was like everything was all about money, and that was…they went into a room together, I was left in another room with my solicitor and we chatted between ourselves and the barrister was coming back and forth, back and forth, asking me more questions about different things and different topics, and then they came back in and said this is the price that we have for you… It was a thing of, here’s some money, go away and shut up now.” [Participant 10]

#### Theme 4: Emotional and Mental Health Impact of Litigation on Plaintiffs

##### Summary of theme

This theme explores the emotional and mental health impact of the decision to pursue litigation, identifying the impact of delays and also of assessments, including invasions of privacy and feelings of violation. Participants identified how these additional burdens are experienced on top of bereavement or dealing with complex medical needs, either of the participant or of their child. Some participants also identified the long-lasting negative impact of these harms on their well-being and mental health.

##### Impact of Delay

Litigation is a protracted process, often lasting for up to 4-5 years. Many participants identified the impact of the delay on their emotional well-being and at times, their mental health and the way in which this protracted process limited their recovery. As described by Participant 3:

> The time frame is absolutely horrendous because I wasted 4 1/2 years of my life…my life was just sitting on hold four and half years of my life. And I suppose I lived in fear as well as looking over my shoulder.

Participant 1 described being consumed by the litigation:

> “Five years. And the big difference was for us, we were consumed by that 24/7 for five years. There was no sense that there was that any similar impact was on the individuals in healthcare, for even on the day of the court hearing, they weren’t present” [Participant 1]

This feeling of wasted time is reiterated by Participant 9 who saw the delays as a deliberate attempt to restrict liability: *“They just, they tried to delay and they tried to break me, they tried to break my spirit.”* Participant 9 continued:

> “So it was very stressful and I really only, I really only understood how much stress I was in when five and a half years later, when I was closed and I woke up the next day with a huge weight off my shoulders.” [Participant 9]

##### Experiences of Assessments

One of the strongest findings to emerge from participant interviews across all contexts was the additional harms imposed by the assessments which were part of the legal process. Participant 6 described how:

> “The assessments, they just went on and on. I had three in one week; they nearly broke me. The things they said… So, these are all disgusting things that I’m so, it took me so long to pick myself up from.” [Participant 6]

Several participants spoke also about the impact of assessments on their children. Participant 15 described:

> “ [I]t was very overwhelming for a child who was kind of between six and eight to have all of this extra going on. Like, these assessments would take between one to three hours. Like, they were horrific”. [Participant 15]

Both the experience of being assessed and the way in which they (or their children) were described in the reports were traumatic for many participants. Some participants found psychological reports especially difficult. Participant 7 describes:

> “I just felt like they were trying to find… obviously they were sending us to these because… and it was only one psychological assessment meeting and you have to obviously go through everything, you know, in great detail. And, like, for them to come out with, you know, how damaged you are or not damaged or, you know. And I have to say for us and for all the other families that had to go through, that was very, very hard” [Participant 7]

The tensions created by ongoing psychological assessments are recognised by Participant 3:

> “But I had to take the kids up to Dublin to see a psychologist for the opposition. Yeah, because they wanted to see how sad they were at their mother is dead, you know?

*And I’m and I’m doing everything I can to make sure these are the happiest kids ever*.

> So now I’m thinking Christ, I might doing too good a job. I’m just gonna mess everything up.” [Participant 3]

Several participants identified how they had been forced to resist violations of their privacy and dignity as part of the assessment process Participant 6 describes:

> “I remember being on a laptop for one, it was an occupational therapy assessment… I was asked to lift the laptop off up the table and pull down my pants and show my lymphedema, and I said to the occupational therapist, if you’re not willing to meet me in person, but you want me to pull down my pants here, I said I am not doing it. Absolutely not.” [Participant 6]

Participant 3 describes the experience of being interviewed about their and their children’s lives:

> “You’re constantly like … a that is just, that’s torture. Like I remember she wanted to go to my bedroom. She wanted to have a look around my house. And I just said – no you’re not going into my bedroom”. [Participant 3]

In circumstances in which participants had already lost trust in the medical system, the way in which assessments operated were sometimes experienced as a deliberate attempt to make their lives more difficult. Participant 14 describes

> “And I suppose it was, like, their indifference to Ciarán. You know, their… you know, not giving a shit, you know, about the way they scheduled the appointments. I was thinking, like, holy god, like, just have a heart, like. But they didn’t seem to care.

> You know, they just kind of… they were hoping… it felt to me like they were hoping that I wouldn’t go for one of those appointments so that they could say, oh, she hasn’t gone to that appointment, we’ll have to postpone it, you know”. [Participant 14]

In addition to the conduct of the assessment, participants identified the negative impact of the way in which the report was formulated. Participant 4 explained:

> “[L]et’s say the urological report, and then you go to somebody from the other side and they’re nice as pie, and they’re, “Oh yes, ok, ah yes, right, yes, no problem’s at all”, and you’re seeing them and you’re telling them what’s going on and all the ailments and everything and all the operations and procedures you’ve had, and they’re nice as pie to your face and you will get a report then, and when you read the report it’s like, WHAT!! But sure…and it just makes you feel that you’ve done wrong”. [Participant 4]

##### Additional burden

All participants had experienced ongoing harms as a result of the original adverse event. Some were bereaved, some were experiencing ongoing physical and/or psychological harms, and some were engaged in supporting children with complex needs. As described by Participant 7,

> “And of course people are broken, damaged, you know, forever grief-stricken after what they’ve been through and after losing their babies.” [Participant 7]

For some participants, the experience of the legal process was more traumatic than the underlying adverse event. As described by Participant 15,

> “Obviously, Samantha’s birth was traumatic and took everything out of me, obviously, and to be left with a child and everything. But the legal proceedings, especially the last two years, were actually worse than Samantha’s birth and I say that very honestly”. [Participant 15]

Similarly, Participant 4, noted that *‘There is so much, you know, and I think the biggest hurt and harm out of all that is how re-triggering and re-traumatising it is, at every step. At every step.’*

The severity of the impact on participants’ lives is evident from several participants. For Participant 6, the negative experience was so severe that they had become suicidal:

> “So, I just got so depressed in the middle of all this. To the point - and I’m being very honest - I nearly ended my life, and I was saved by a woman and dog at a beach, yes. And to this day, I can see her. It had just wrecked my whole being”.[Participant 6]

##### Ongoing mental health/well-being difficulties

Several participants indicated that they continued to have mental health/well-being difficulties which in some cases continued long after the legal process had concluded. Participant 6 describes leaving the court and almost walking under a bus and explained that *‘It took me months, months for my body to regulate again, my sleep to come back, my bowel was hugely affected and plus all the other side effects with it.’*

##### Participant 12 explained

> But I literally, if I walk into a hospital, I might have a panic attack. I have… I do have nightmares. I do have really bad panic attacks. Now, I’m doing better with those now because I started counselling. But it’s taken a long, long time to get there because I felt that if somebody in the medical profession could lie to you like that, then what hope is there for anybody. [Participant 12]

#### Theme 5: Advocating for Change: Participant Recommendations

##### Summary of Theme

Participants provided valuable insights and recommendations to improve the process following patient safety incidents and navigating the litigation route. This theme encapsulates their diverse recommendations, reflecting their experiences and perspectives and a collective effort to improve the experience of plaintiffs, promote transparency, and foster a more supportive and effective system for addressing patient safety incidents, patient and family wellbeing and litigation.

##### Key Recommendations

1. Open Disclosure: A key recommendation from participants was the need for the implementation of Open Disclosure as part of patient and family support following a patient safety incident.

> “Open disclosure, for a start. Because there was none of that in our situation, in any way, shape or form. Just, as I said, open disclosure. Sit down and actually have a conversation with these people and find out what exactly happened. ..You know, I know for a fact we would have walked away and just… this is what happened” [Participant 7] The importance of implementing Open Disclosure as a necessary step in the duty of care and not just as a mechanism to avoid litigation was recommended.

> “When you’re doing open disclosure or any of that..you can’t be doing it with something at the back of your mind. If I do this right, they won’t litigate, they’re entitled to the information. What they do with the information afterwards is no concern of yours. They may decide to go for litigation and that is their right, and you can’t blame them for that and you can’t think less of them for that.” [Participant 1] An integral step to the implementation of Open Disclosure is the need for change in culture within healthcare institutions so that staff feel supported to implement Open Disclosure and to engage in open and honest communication

> “I thought that [Open Disclosure] would have made a difference, but it doesn’t seem to have made that much of a difference because you still get, you know, horrible stories… I find it incredible that there still is a culture of hiding the truth, you know, and umm, that’s what I would change… We understand that that’s an instinctive thing, but you know, we also understand that, you know, we need the the environment needs to exist to allow a person to go back and say, look, I could have got it wrong. Here’s what I think might have happened. You know, together with all the the the support of psychological safety that’s been put in place. And yeah, I do despair that despite all that, you still hear stories.” [Participant 2] This change in culture also requires collaborative communication, which promotes collaborative discussions and listening to patients/families to address needs and avoid adversarial dynamics.

> “So they could have eased all of that and they could have had, you know, sat down properly and realised as well, ok look lads, we’re at fault here really to be fair like. You know what I mean. So, we’ll just – let’s go talk to them and let’s, you know, let’s sit down and, you know, plough something out. But, they didn’t and they fought us really, basically they fought us all the way” [Participant 4]
2. Mediation and Alternative Pathways: Advocating for mediation and alternative dispute resolution methods at an early stage.

> “I think mediation because people are, you know you’re dealing with broke; you’re beaten, you’re dealing with broken people on both sides. You have to, and a safe place to, a safe place to express your expressions and not, not lose that communication, not break that strand of communication.” [Participant 5] Key to the success of this approach is ensuring that mediations are not simply settlement talks facilitated by a third party, rather they should provide an opportunity for parties to collaboratively explore and negotiate mutually acceptable resolutions to their disputes.

> “even with the mediation, kind of, the HSE were in one room, our legal team were in one room and we were in one room. So while we were supposed to be the centre of it, we were still very much on the outside of it. You know, which *was very difficult because, like, our own legal team were in the room for four hours and didn’t come to even speak to us, do you know. So, like, this was about us and about us having mediation and we were very much, I suppose, bystanders and spectators in this.”[Participant 15]*
3. Empowering Plaintiffs: Participants stressed the importance of involving litigants in the process and ensuring their voices are heard to prevent them from feeling sidelined or lost within the system.

> “I think I think there needs to be more involvement of the litigant in in that as well. How that would look like, I don’t honestly know but it was this sense of that it’s out of your hands, and because he was so precious to us, and because the whole incident was so appalling, I know I for one, didn’t like this feeling of being sidelined because part of our healing was our doing our bit of the job and when we hit this point [litigation], it looked like we had no role to play. And it was that bit again that we were kind of at the mercy of yet another system” [Participant 1]

> “..for people, men and women like myself that have been affected by negligence and has had to go through the legal system is…to be able to sit down in front of a legal system say…jury if you like it, and explain the damages. I think they need to hear it from the ground first before it, because I know like, they’re just doing their job and they have protocol and they have a system and everything, but the person gets lost, the real person gets lost in it. I got lost in this as a person, I was just case number hm-m-m” [Participant 6]
4. Providing emotional and psychological support to patients and their families following an adverse event.

> “No sense of listen, you’ve had a big trauma, do you need a bit of support? A bit of help? … this is going to have a big impact on you. Already, I can see your, you know… You should check in with the psychiatry team there because it’s… Not today, but you’re gonna need a link. And and I didn’t and but I needed somebody to say, look, you’re going to need support here and nobody did. There was no holistic care of any kind.” [Participant 11]

> “they need to sit down and go is look, this is what we think this means. Let’s have an ongoing conversation. You know, sit down with a counselor, sit down with a pediatrician, sit down with people who understand what the trajectory of this is. This is going to have lifestyle implications in different ways. You’re going to need care.” [Participant 11]
5. Providing early support and care to babies leaving hospital to ensure they receive necessary support at early stages:

> “… I needed medical support when Samantha was born and brought home. I needed therapeutic support when I brought Samantha home. Like, I… I suppose I read everything, I went to seminars, I did everything. I literally trained myself up to be an OT, SLT, physio, nurse, doctor, you name it, do you know, off my own back. Whereas if I had those people when Samantha was born… it would have helped us an awful amount. First of all, for Samantha, second of all for ourselves.” [Participant 15]
6. Support during litigation: Participants advocated for providing non-legal support to plaintiffs during litigation to alleviate vulnerability.

> “There should be another person that meets with the victims, gets to know them, has a chat and is present with them. Not give you legal advice because that’s not their duty, but do you understand what I’m saying, just to take the edge out of it because you are so vulnerable.” [Participant 6]
7. Streamlining Expert Assessment Processes: The necessity to implement changes in assessment procedures to minimise repetition, speed up evaluations, and reduce the burden and the trauma caused.

> “Yes, I had to do double. I had to get all my reports, my psychological reports, nurse caring reports, I had to get all those reports done a second time because, Oh so much time went by, they wanted updated ones. And in the end, in the end, it didn’t matter.” (Participant 9)

> “I think when they’re doing the reports, so when they’re doing the expert witness reports, could not, you know, the HSE and the legal, our own legal team have the two, say, OTs sit in one room and assess her at the same time, to speed it up a bit besides doing the same assessment on two different days in two different places. I think that’s firstly” (Participant 15)
8. Efficiency and expediency in the litigation process to prevent prolonged distress

> “So there has to be a more humane, you know, patient-centered approach to this and that has to be efficient and effective in a timely manner also…The time frame is absolutely horrendous because I wasted 4 1/2 years of my life… my life was just sitting on hold four and half years of my life. And I suppose I lived in fear as well as looking over my shoulder. Constantly being careful what I said. Being careful what I did because I knew I was constantly being watched as well” [Participant 3]

> “The Defendants need to be put on trial for stealing five and a half years of my life, because they’ve wasted, they wasted so much time. It was dirty, dirty tactics. They just, they tried to delay and they tried to break me, they tried to break my spirit.” [Participant 9]

> “Five years. And the big difference was for us, we were consumed by that 24/7 for five years. There was no sense that there was that any similar impact was on the individuals in healthcare, for right up to the day of the court, they weren’t present” [Participant 1]
9. Strategy following Admission of Liability: Calling for quicker resolution processes in cases where liability is admitted, to avoid prolonged litigation and unnecessary court involvement.

> “It should have been a case of, you know, “Yes, we’ve screwed up. Here’s your letter. We’re really sorry and here’s what we’re going to give you for it” [Participant 8].

> I just think when something is such clear cut. Two people had said yes, we’ve made a mistake, you know… Court, I just don’t think we should have been there. Just plain and simple, I just think it was an awful waste of people’s time, it was an awful waste of public money. And it was just a bit of a joke, like.. Yes, my big bugbear was I just thought it was an awful waste of money, us being in court for three days. *[Participant 14]*
10. Reviewing the statute of limitations for parents taking cases related to a birth injury.

> “the two years, first of all, the two years needs to change. It needs to absolutely – a birth injury for two years, you’re not even in your sane self. Even a person who has a normal delivery…as a mother, you’re not yourself. You’re not yourself, your sleep and everything. So I think that is a huge factor in, I think that’s a huge factor in post-natal depression long term, in that if something has happened, you don’t have time down the road to deal with it. So that’s probably one thing that I’d love to see maybe implemented or changed. You have the right to fight your case, you know, when you’re ready. You know, whether that’s you putting in an Affidavit saying, right, “Can we just pause a few years”, and then let me come back to it in a few years.“ [Participant 13]
11. Governance and Accountability: Emphasizing the importance of monitoring occurrence and reporting adverse events and tracking these against litigated cases in hospitals to hold hospitals to account and prevent recurrence.

> “where’s your responsibility in checking the national incident management system to make sure that everything has been reported and have have you got litigated cases that weren’t reported on the incident management system? And what’s your feedback loop to hold hospitals to account?” [Participant 11]

> “How many incidents are referred to the National Incident Management System, per hospital, per maternity hospital. And do you then verify it? So for example, was my son’s case on the National Incident Management System? Of the cases that turned into litigation do you look back and see if they were verified or not? Because if they’re not, then that’s some ********. Where is the accountability with that? And then do you go back and look at the types of cases that are coming out and do you have a pattern of cases in relation to

> [X] in [Y hospital], for example, or something like that? Or is there a pattern where…And and how do you go back and test thisOr when are the days that you bring the hospital management in and hold them to account for the different events that have occurred? That the state is now liable for. Because you’re not doing it, are you?” [Participant 11].
12. Advocacy for Women’s Health Equity: a strong call for action to address the systemic issues affecting women’s health in Ireland, advocating for greater gender equity, empathy, and respect within the healthcare system

> “So I think the way we treat women in this country medically needs to be changed. I would change how the medical profession and… treat women. I would change how we refer to women in documents, particularly mothers. Particularly first-time mothers. Any mother.” [Participant 12]

### 1.3.4. Submissions to the Expert Group to Review the Law of Torts and the Management of Clinical Negligence Claims

The submissions made to the Expert Group [38] provide a further source of direct evidence of the plaintiff experience and allow for a degree of triangulation of the findings of the qualitative description study. There is a notable consistency between the submissions to the Expert Group and the findings presented above.

The key finding that litigation is a last resort, taken only when other options are unavailable, is entirely in accordance with evidence provided by Sage Patient Advocacy. In their submission, Sage stated:

> From our vast experience in providing a patient advocacy service it is our view that for the majority of people the process of seeking redress through the legal system is a last resort, and a decision taken at a time when the person feels they have no other option as the other avenues through the established complaints and review process have been exhausted without receiving a satisfactory response.

Sage identified that:

> The decision to pursue a case through the Court system is taken with careful consideration and primarily in an effort to establish the facts and to be given an explanation by the healthcare system and the healthcare personnel involved in their care or their loved one’s care.

The Sage submission also referred to three recent cases in which the organisation had been involved which they present as evidence that alternative ways of dealing with people who have suffered harm may help to avoid litigation:

> We have recently been involved in three cases which were resolved through an open disclosure process, in two of these cases the people affected who were seeking an explanation did not pursue a legal route as the healthcare provider engaged on an equal basis with the person, acted in a transparent manner, provided the facts and an explanation, and provided an apology. In the third case the patient had already engaged with a solicitor prior to the open disclosure meeting taking place. In this case the family had to request a meeting with the hospital to try to get explanations as to what happened.

The submissions also support the finding that the legal process feels imbalanced to plaintiffs. The mother of a daughter born with cerebral palsy identified the issues of costs, an issue which also emerged in the case study:

> The cost to me as an ordinary parent to take a court action on behalf of my…daughter is prohibitive, .. the cost of providing expert witness both medical and legal is far too expensive for the average parent and as the State can call on many of these at my tax expense.

A consultant medical oncologist identified that the problems resulting from litigation are experienced by both patient/family members and professionals stating:

> The adversarial nature of the current medicolegal structure worsens patient and family upset and the protracted nature of these cases as currently conducted worsens health care provider anxiety.

The submissions also support the claims made regarding the systemic nature of the problems identified. The consultant medical oncologist identified the absence of any opportunity for learning from the court process, noting the ‘lack of any feedback learning system for clinicians of medicolegal claims’. He describes a lawsuit in which he was involved which had settled and notes ‘no feedback was given to doctors involved of how practice could change nor was there discussion of the valid claims by the plaintiffs medicolegal advisors about how care could be improved.’ He concludes ‘[i]f we are not aware of analysis of our mistakes we will be condemned to repeat them’.

## 1.4 Discussion and Conclusion

It can be seen from the literature and the findings of this research that many patients, subject to injuries resulting from medical negligence, find themselves subsequently re-victimised by the often long-drawn-out, and over-bureaucratic legal forums.

The Review of the Implementation of Recommendations of the Scoping Inquiry into the Cervical Check Screening Programme published in November 2022 [39] highlighted the rights of patients to know the truth about their health.

I. deserve to know the truth To be told the truth
II. *I deserve answers I deserve justice*
III. *I deserve for this never to have happened I deserve closure*
IV. *I deserve for it to be over*,

[I Deserve - compiled from the words of 221+ members by artists Fiona Whelan and John Conway.]

These words, the experiences presented in this research, and the evidence from the literature, highlight how actions taken in the aftermath of a patient safety event have far-reaching consequences for patients, their families, healthcare professionals, and the organizations involved. Liukka and colleagues conducted a review of the literature on actions after an adverse event and categorised these actions based on different stakeholders [40]. Common elements across all stakeholders include the imperative for empathic and ethical communication, provision of support services, issuing a complete apology, and facilitating training and learning. Specifically focusing on patients and their families, these actions encompass the disclosure of the adverse event, communication post-event, provision of emotional and compensation support, and the delivery of a comprehensive apology to address the concerns of patients and their families [40]. These elements align to the HSE’s Principles of Open Disclosure which include acknowledgement of the incident, truthfulness, timeliness and clarity of communication, apology/expression of regret, and recognising patient and carer expectations.

Deficiencies in the delivery of these actions were identified by participants when discussing their interactions with healthcare professionals and healthcare organisations in the immediate aftermath of the event and in the days, weeks and months following. Patients and their families often have a sense of being wronged and this is compounded by a situation of overt and perceived cover-up, defensive staff encounters and use of language which is insensitive and sometimes antagonistic. Staff involved in these encounters can have a lasting impact, both positive and negative, on patients and their families. Participants in our study very vividly described their encounters with different individuals much more than the organisations involved. It is essential that organisations support their staff through appropriate implementation of policies and procedures and provision of staff training to ensure that ‘on the ground’, open disclosure is practiced. Organisations should also recognise the importance of providing reassurances to staff about their continued support should the litigation route be engaged. By fostering a culture of openness, empathy, and learning from mistakes, the practice of open disclosure has the potential to negate the need for litigation. The recently published National Open Disclosure Framework (2023) aims to provide a unified and consistent approach to open disclosure across the sector and places responsibility on each organisation to adopt the Framework and to “embed positive open disclosure cultures and behaviours into practice”.[41] It is this obligation to incorporate practices and behaviours which promote a culture of open disclosure in practice and the monitoring and evaluation steps contained therein which will be of great significance in the future.

While financial compensation is a need for many, other factors carried an equal if not greater significance to the patients. These included, an explanation, an apology from the clinician/physician responsible, an opportunity to be heard, assurances that recurrence of such negligent conduct would not befall other patients, practical supports and suitable emotional redress tailored to their specific needs. As outlined in our research, money is often not the motivating factor and pursuing the litigation route is often unsatisfactory in achieving these ‘extra-legal’ goals; too much time has passed, apologies if received, may seem ‘too little too late’, and the healthcare professionals involved are excluded from the system. When financial compensation is sought, participants emphasized the need for a patient-centred approach to resolving their case, particularly so when extensive treatments and supports are required following patient safety events. Instead of resorting to litigation, which can be time-consuming, adversarial, and emotionally draining, participants advocated for more expedient and collaborative methods such as mediation.

The experiences of patients and families of the medico-legal environment is arduous, lengthy, frustrating and described by a participant in our case study as a “David and Goliath” experience. A barrister who participated in a study on medico-legal actions in Ireland published in 2021 summarized this point “Don’t ever think for a second, going to court is ever going to be, in any adversarial setting, easy” [8]. The protagonist of the litigation process is the legal profession and several participants in our study described a feeling of being ‘side-lined’ in the process. Mediation, as an alternative dispute mechanism, is suited to many clinical negligence claims and is identified as the “default position” of the States Claim Agency in how they prefer cases to be handled [42]. The Expert Group to Review the Law of Torts and the Management of Clinical Negligence Claims outlined that as part of any reform, mediation should be established as a part of the process. The Mediation Act 2017 places an obligation on legal professionals to inform clients about mediation as a potential route to pursue prior to issuing proceedings, but it is acknowledged that mediation has been underused for cases of medical negligence [8]. An exploration of the use of mediation in medical negligence claims in Ireland by Mitchell suggests that mediation when used, is done as part of “the convoluted litigation system and the style used is focused on the legal interests of the parties rather than any emotional needs” [43]. This is supported by this research which evidences the fact that mediations, as currently conducted in this jurisdiction, appear to be akin to settlement talks facilitated by a third party, dominated by the legal teams with little involvement of the parties involved. If mediation is to reach its full potential and to provide the benefits anticipated in the Report to Review the Law of Torts and the Management of Clinical Negligence Claim, including meeting the needs of plaintiff/patient for full information about what happened, it will be necessary to ensure that mediations are conducted differently. The plaintiff must be at the centre of the process and mediations focused on meeting the aims, including the ‘extra-legal’ aims, of those involved.

This research has evidenced a lack of understanding amongst participants about their options following an adverse event, with participants being pointed towards the legal profession both formally and informally in their quest for information, understanding and resolution. This research also evidenced a lack of understanding amongst participants about how their case was resolved. This points to an opportunity to engage with and empower patients and their families at the earliest opportunity following an adverse event and to provide them with opportunities to meet their needs without recourse to the legal process or should they find themselves in a litigated process, less vulnerable because of a lack of knowledge and understanding. Mitchell proposes that implementing a public educational program focused on presenting alternatives to litigation, such as mediation, could enhance awareness and promote greater involvement in restorative resolution approaches [43].

The findings of this research underscore the clear advantages of implementing the objectives and actions outlined in Goal 2 of The Justice Plan 2023 [44], particularly those concerning medical negligence cases. Of particular relevance are the establishment of the Mediation Council of Ireland and introduction of Pre-Action Protocols. While case management and pre-action protocols were not addressed directly by our participants, this is unsurprising given that they would have no direct experience of either. However, it is clear that both mechanisms provide an opportunity to address some of the concerns raised in this research, particularly to limit the damage caused to plaintiffs by excessive delays.

The studies reviewed included jurisdictions which operate in a similar fashion to Ireland (e.g. United Kingdom), and countries which have vastly different approaches (e.g. New Zealand, Norway or Netherlands), each one providing insights from patients with common concerns and desires for openness and transparency following any wrongdoing. It can be seen that any reforms in the future must be aligned with patient needs, and that lessons can be learned from any transgressions in the past, such as avoiding defensive practices by physicians, and engaging with patients in a more ethical manner from the identification of the error in order for the patient’s voice to be heard and inform the process.[45]

> “Let the young know they will never find a more interesting, more instructive book than the patient himself.” (per Giorgio Baglivi 1668-1707)

### Strengths and Limitations

In terms of credibility, dependability and transferability, i.e., trustworthiness, of this research, we must be cognisant of the assumptions of our heterogeneous research team influencing our analytical interpretations. Three team members have a legal background and three are from the health professions (pharmacy, nursing, medicine). All are higher education faculty members no longer working in clinical (legal or healthcare) practice. As qualitative researchers, all have experience in thematic analysis. Given this, we as researchers understand that

> qualitative researchers seek to understand the world from the perspectives of those in it. Since there are many perspectives, and many possible interpretations…Instead of reliability, we can strive for…consistency and dependability…whether the results of the study are consistent with the data collected [46]

Therefore, to strengthen the internal validity of this research we familiarised ourselves with the data again and again returning to the text and sharing our interpretations, using reflexivity to ensure extensiveness of data analysis. We triangulated our findings with similar cases in the literature, inclusive of legal and policy document analysis. We engaged in member checking: by sharing our findings with our participants to check for congruity.

A significant strength of this study includes its in-depth exploration of participants’ perspectives and experiences regarding patient safety incidents and litigation. The use of qualitative methods allowed for a nuanced understanding of the phenomenon of medical negligence and its impact on patients and families in the aftermath of the event and subsequently through the legal system. Additionally, the study employed a diverse sample of 15 participants geographically spread across Ireland, capturing a range of voices and experiences, thereby enhancing the credibility and transferability of the findings. In addition, the inclusion of five participants who had experience of a patient safety event at birth, or the neonatal period adds to the existing evidence-base and is particularly important given that catastrophic birth injuries account for a significant proportion of medical negligence claims in Ireland. However, it should be recognised that those participants who volunteered for this research have a strong level of commitment to change and so we should be clear to adhere to the concept of transferability rather than generalization of these study findings.

This study sheds light on the experiences of patients and families following a patient safety event. The imperative for Open Disclosure is highlighted not only to address the ‘extra- legal’ requirements of explanation, an apology, and the prevention of the injury happening again but also to ensure accountability within the healthcare system. The intricate dynamics between financial compensation, justice-seeking, and the healthcare and legal systems are discussed and contribute valuable insights to the discourse on medical negligence in Ireland.

## Data Availability

The participants of this study did not give written consent for their data to be shared publicly, so due to the sensitive nature of the research supporting data is not available

## Acknowledgements

The researchers would like to sincerely thank all the participants in this study for sharing so openly their stories. We are very grateful for your invaluable contribution to the research.

We would like to thank the staff in the National Quality and Patient Safety Directorate and the National Open Disclosure Office for facilitating recruitment to the study.

